# DREAM: A framework for discovering mechanisms underlying AI prediction of protected attributes

**DOI:** 10.1101/2024.04.09.24305289

**Authors:** Soham U. Gadgil, Alex J. DeGrave, Joseph D. Janizek, Sonnet Xu, Lotanna Nwandu, Fonette Fonjungo, Su-In Lee, Roxana Daneshjou

## Abstract

Recent advances in Artificial Intelligence (AI) have started disrupting the healthcare industry, especially medical imaging, and AI devices are increasingly being deployed into clinical practice. Such classifiers have previously demonstrated the ability to discern a range of protected demographic attributes (like race, age, sex) from medical images with unexpectedly high performance, a sensitive task which is difficult even for trained physicians. In this study, we motivate and introduce a general explainable AI (XAI) framework called DREAM (DiscoveRing and Explaining AI Mechanisms) for interpreting how AI models trained on medical images predict protected attributes. Focusing on two modalities, radiology and dermatology, we are successfully able to train high-performing classifiers for predicting race from chest x-rays (ROC-AUC score of ∼0.96) and sex from dermoscopic lesions (ROC-AUC score of ∼0.78). We highlight how incorrect use of these demographic shortcuts can have a detrimental effect on the performance of a clinically relevant downstream task like disease diagnosis under a domain shift. Further, we employ various XAI techniques to identify specific signals which can be leveraged to predict sex. Finally, we propose a technique, which we call’removal via balancing’, to quantify how much a signal contributes to the classification performance. Using this technique and the signals identified, we are able to explain ∼15% of the total performance for radiology and ∼42% of the total performance for dermatology. We envision DREAM to be broadly applicable to other modalities and demographic attributes. This analysis not only underscores the importance of cautious AI application in healthcare but also opens avenues for improving the transparency and reliability of AI-driven diagnostic tools.

## Introduction

Image-based or visual tasks are common in medical practice, and artificial intelligence (AI) has been used increasingly for these tasks. In fact, the United States Food and Drug Administration (FDA) has cleared hundreds of AI devices for medical image classification tasks, with more devices still under development^1–3^. As AI becomes embedded in clinical decision-making processes, ensuring the robustness and fairness of these models must become an integral part of their development. Exposing these mechanisms offers transparency into the reasoning processes of the AI models and provides developers with the opportunity to reduce model vulnerabilities that could lead to discriminatory performance in downstream clinical tasks.

To generate their predictions, AI models not only rely on medically reliable image attributes, but also sometimes rely on irrelevant and likely undesirable attributes like image acquisition artifacts^4,5^ that can degrade a model’s predictive performance. Evidence has also shown that AI-based medical image classifiers may leverage protected demographic information to generate predictions;^6,7^ inappropriate use of such attributes (e.g., race or sex) could lead to fragile predictive performance or discrimination due to domain shift in clinically relevant tasks. Perhaps more concerning, AI models have unexpectedly displayed the ability to predict a range of demographic variables directly from medical images^8–10^ even in the absence of demographic metadata. Examples include the prediction of a patient’s sex from retinal fundus images^8^ and the prediction of a patient’s race from different forms of radiological imaging.^9,10^ If these demographic variables correlate with a diagnosis or prediction target in the classifier’s training data—either due to societal inequities or random chance—an AI model that uses all available information is likely to incorporate these variables in its prediction. Prior work reveals that such classifiers can use these demographic shortcuts in disease classification, leading to biased predictions across subpopulations and fairness disparities across both in-distribution and external test sets.^9,11^ Significantly, physicians in relevant imaging fields cannot explain how AI models are able to predict certain demographic features from medical images. This paper proposes DREAM (DiscoveRing and Explaining AI Mechanisms), a general framework to examine and quantify these unknown signals underlying AI predictions of protected attributes from medical images in a human-interpretable manner.

Prior work attempting to tackle this problem has only partially explained the high accuracy of these classifiers,^9,12–15^ citing factors like correlations between protected demographic variables and diagnoses or other variables more visible in the images (e.g., age). However, it fails to quantify the degree to which each signal contributes to overall classification performance. Prior work has also been tailored toward specific modalities and does not provide a generalizable approach. Further, the unexplained performance of these AI models has led to further speculation about the existence of specific image signals which are leveraged by the models to make these predictions.

The postulated existence of AI-specific signals raises two questions that we also address: (1) *How do medical image classifiers ‘reason’ ?* That is, what signals do they use to generate predictions and to what extent? (2) *How might such classifiers produce disparate outcomes among protected classes*? By applying the systematic approach in our DREAM framework to two different modalities, we study the existence and nature of visual signals that could be used to predict a protected demographic variable, and address a gap in our knowledge of how medical AI classifiers generate predictions, which currently includes only unquantified image attributes readily recognized by humans.^5^ Simultaneously, we detail a mechanism by which classifiers might display undesirable characteristics across protected classes.

Using DREAM, we examine two medical image classifiers trained across two modalities – one to predict sex from dermoscopic lesions, which offer a magnified view of a patient’s skin lesion, and the other to predict race from chest x-rays. To motivate our analysis, we first design an experiment which highlights the detrimental effects of demographic encodings on clinically relevant downstream tasks. Then, we apply a range of methodologies from the field of explainable AI, namely, *clustering analysis* and *counterfactual image generation* to identify signals that may be leveraged by the classifiers for their predictions. Finally, we introduce a novel technique called *removal via balancing*, based on propensity score reweighting, that quantifies the importance of each identified signal to the overall prediction. Figure 1 summarizes our investigative DREAM framework.

**Fig. 1.**
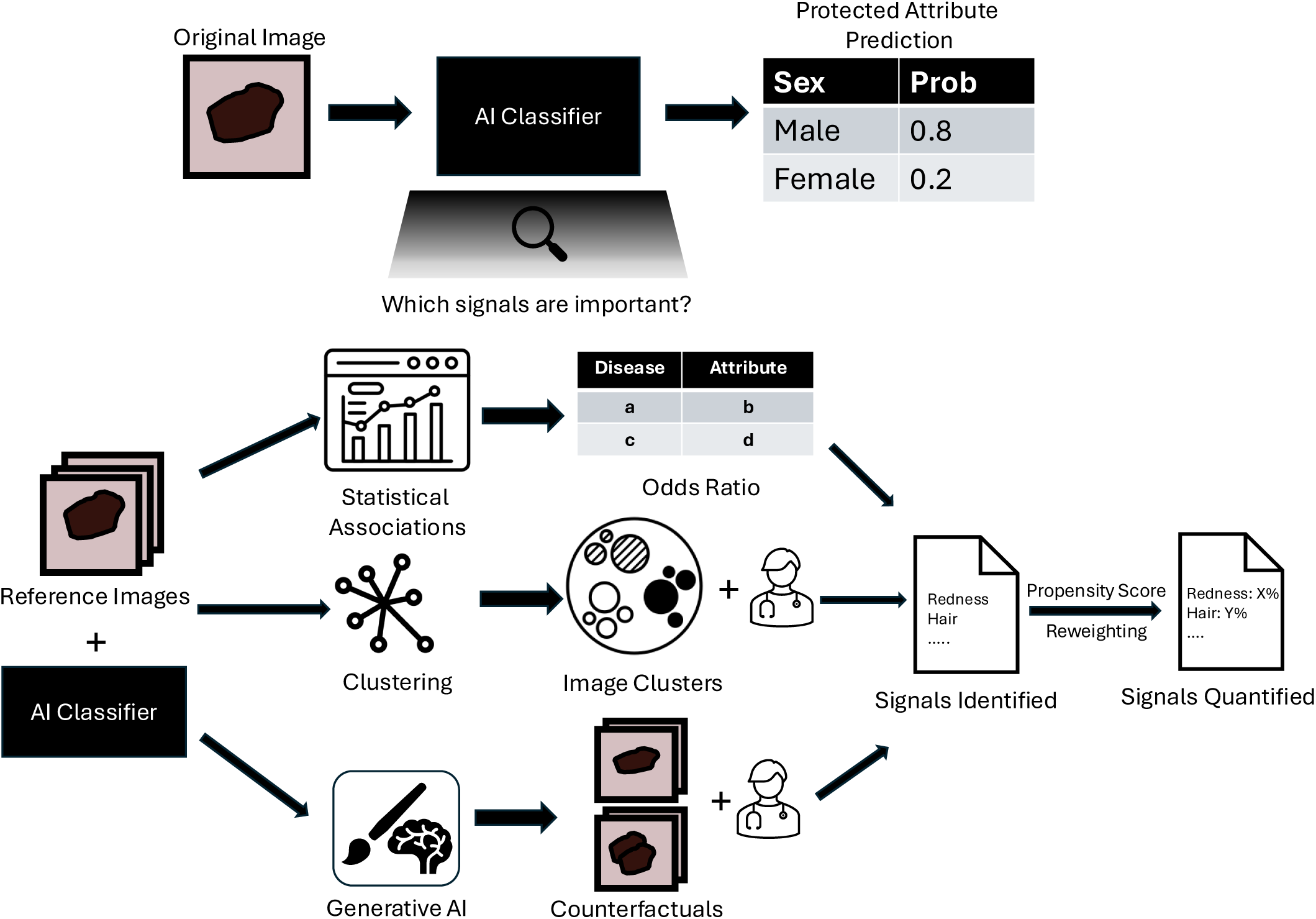
DREAM Framework overview. We first trained a ViT-based AI classifier to predict the protected attribute. Then, using statistical associations and the explainable AI techniques of clustering analysis (for global artifacts) and counterfactual generative AI (for local artifacts) with expert analysis, we identified signals of potential importance to classifier predictions by leveraging the expertise of medical experts. Finally, using our *removal via balancing* technique based on propensity score reweighting, we quantified the importance of each identified signal to the overall prediction. Dermoscopic lesions are shown here for illustration purposes, the same analysis was replicated for chest x-rays.

To our knowledge, we are the first to propose a framework that can be used for rigorous model auditing to (1) identify specific signals that contribute to the predictive performance of protected attributes from multiple modalities and (2) quantify the extent of the contribution. Although this study focused on sex prediction from dermoscopic images and race prediction from chest x-rays, DREAM is flexible and can be systematically applied to other modalities (e.g., retinal fundus images) and protected attributes (e.g., age) that we anticipate could reveal similar vulnerabilities.

## Results

### AI classifiers successfully predict protected attributes

To investigate how AI models predict protected attributes from medical images, we trained classifiers for the specific tasks of identifying a patient’s sex (male or female) based on a dermoscopic image of a skin lesion (Figure 2a) and identifying a patient’s race (black or white) based on a chest x-ray scan (Figure 2c).

**Fig. 2.**
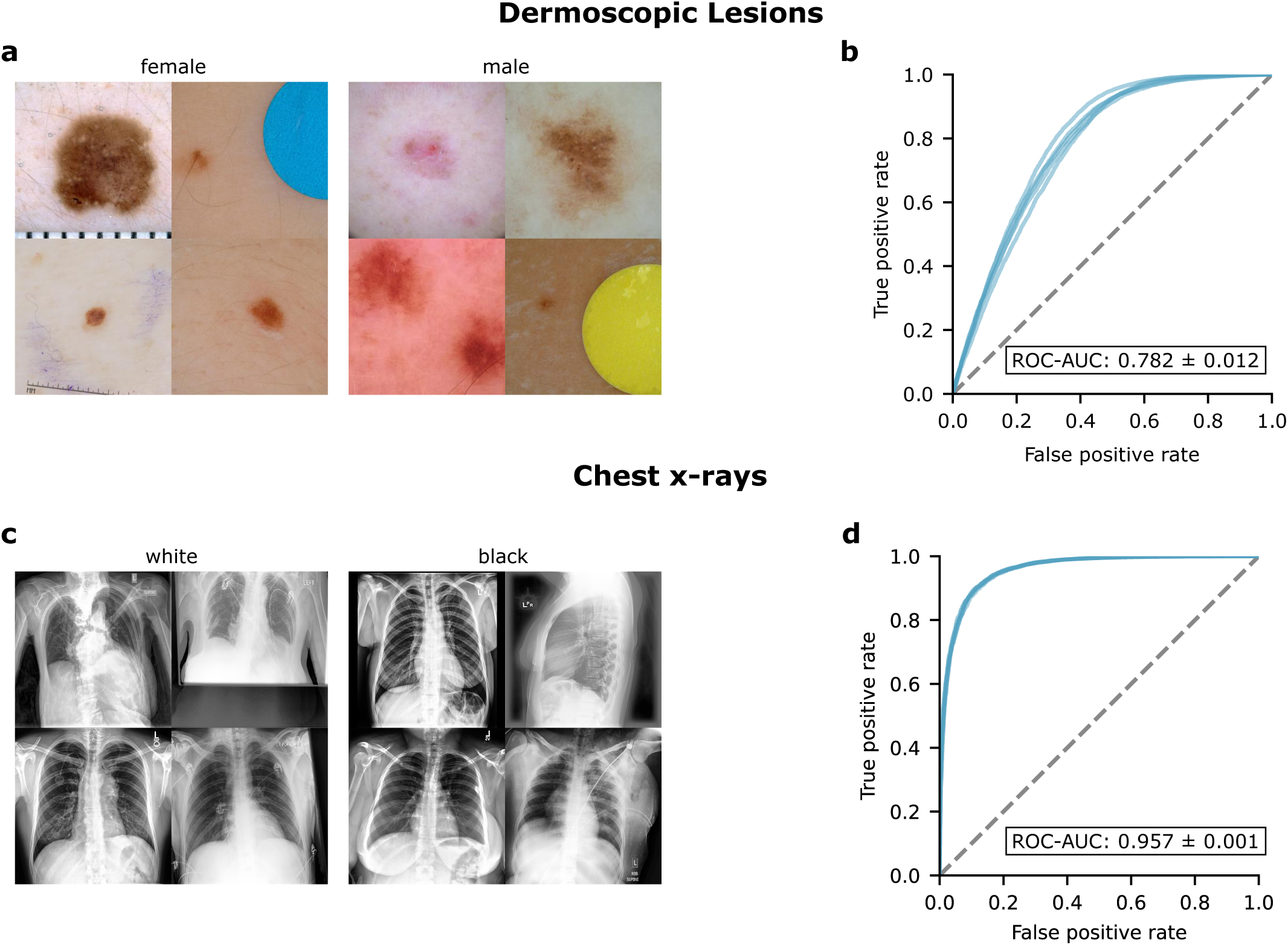
AI prediction of protected attributes. **(a)** A randomly selected set of dermoscopic images from female and male patients. **(b)** Performance of trained AI classifiers, based on a vision transformer architecture, at prediction of sex from dermoscopic images across five replicates. **(c)** A randomly selected set of chest x-rays from white and black patients. **(d)** Performance of trained AI classifiers, based on a vision transformer architecture, at prediction of race from chest x-rays across five replicates.

For the dermoscopic images, we trained our classifiers on images from the International Skin Imaging Collaboration (ISIC) archive. To mitigate the possibility that the model would rely on source-specific confounding rather than signals that generalize across data sources, we partitioned the ISIC archive based on the image collection site and used a disjoint group of collection sites for training and testing. In this external test scenario, our classifiers predicted patient sex with substantial performance (area under the receiver operating characteristic curve, ROC-AUC, of 0.782 ± 0.012, mean ± standard deviation; Figure 2b).

For chest x-rays, we trained and tested our classifiers on different datasets (CheXpert^16^ and MIMIC-CXR^17^) collected from two different hospital sites to reduce source-specific confounding. Even in this scenario, the classifiers were able to predict patient race with unexpectedly high performance (area under the receiver operating characteristic curve, ROC-AUC, of 0.957 ± 0.001, mean ± standard deviation; Figure 2d).

### Prediction of protected features enables undesirable outcomes

Although we typically would not expect medical image classifiers to be used to predict protected attributes, we hypothesized that their ability to do so could lead to undesirable behavior in medically related prediction tasks. If true, this hypothesis would motivate the need for a framework to understand the mechanisms that underlie the classifier’s prediction of protected attributes.

We conjectured that one mechanism by which AI prediction of protected attributes may degrade performance at medically relevant tasks is if the classifier learns to use the protected attribute as a ‘shortcut’^18^ for identification of a disease due to *an association between the attribute and disease in the training data*. Although some associations could reflect genuine medical differences (e.g., rates of breast cancer among females and males),^19^ they could also reflect societal disparities or other spurious variations. If the association changes—or, in the worst case, *reverses*—at test-time (e.g., deployment), then a model that learns to leverage this association from the training data will likely decline in performance.

To test our hypothesis, we focused on the tasks of differentiating melanoma from look-alike lesions (benign nevi, seborrheic keratoses, solar lentigo, etc.; see Methods) and of predicting pleural effusion from chest x-rays. We en-gineered datasets to exhibit an association between the protected attribute and the disease target (Figure 3a). We conducted our tests for a variety of odds ratios, in each case setting the ratio in the external test data to the inverse of that in the training data.

**Fig. 3.**
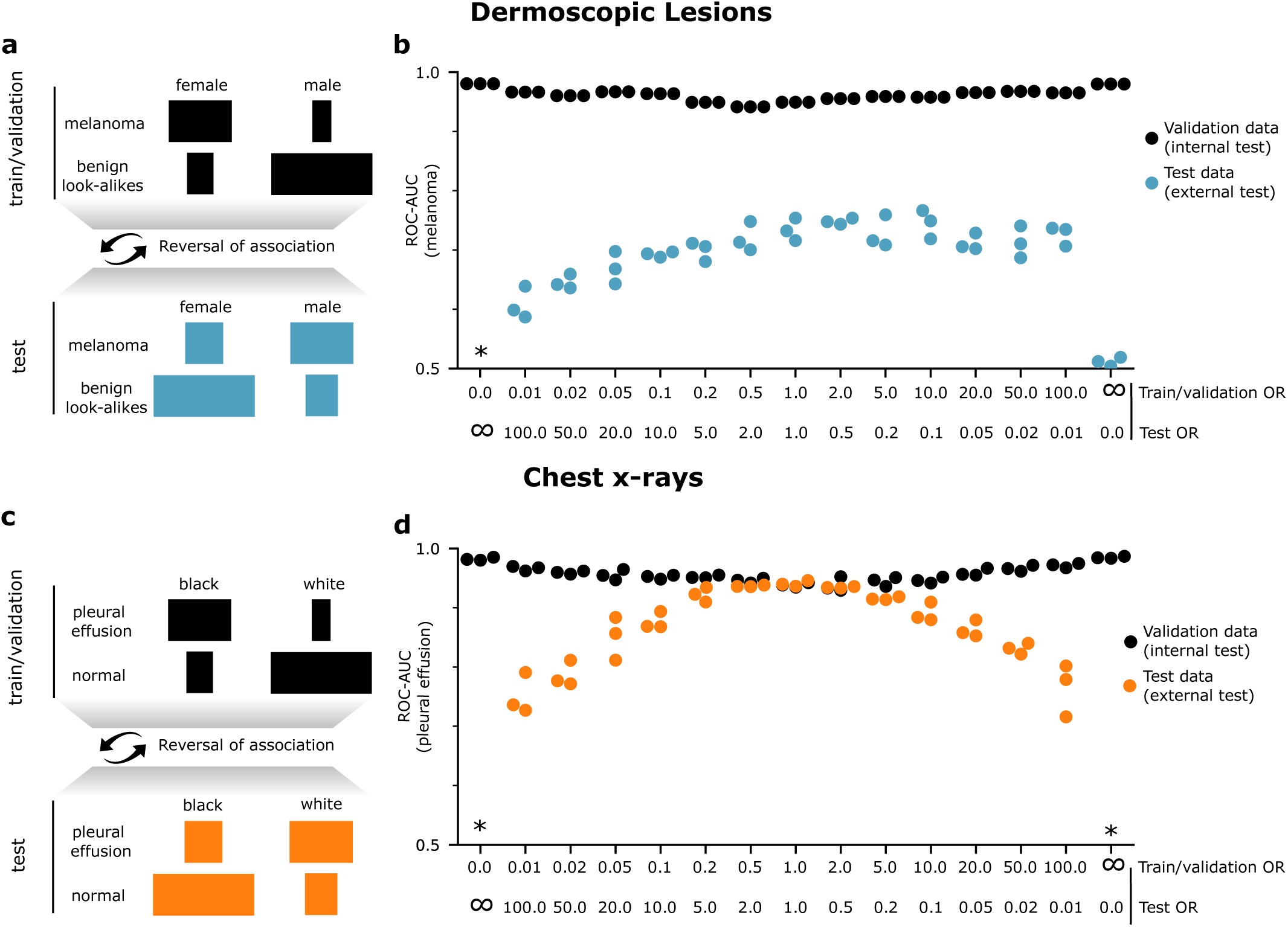
Spurious correlation with a protected attribute may result in performance deficit. (a, **c)** Overview of the experiment’s setup. We engineered datasets (by sub-sampling the original data) such that the prediction target (melanoma from dermoscopic lesions or pleural effusion from chest x-rays) correlated with a protected attribute (patient sex or race). The correlation was inverted in the test data. **(b, d)** Performance of the trained classifiers on the melanoma and pleural effusion prediction tasks across three replicates. *Odds ratio (OR)* measures the association between the demographic ‘female’ or ‘black’ and the prediction target ‘melanoma’ or ‘pleural effusion,’ that is, an odds ratio greater than one indicated that a higher proportion of the corresponding protected attribute depicted the disease. An asterisk (^∗^) indicates ROC-AUC< 0.5 (that is, worse than random performance).

We observed that for both melanoma and pleural effusion, although performance on internal validation data remained high across scenarios (indeed, even improving with stronger associations between the protected attribute and the target), external test set performance declined as the odds ratios varied from unity (Figure 3b). Performance dropped precipitously for extreme changes in odds ratios (in particular, when the protected attribute correlated perfectly with the prediction target). Performance changed more modestly for moderate differences in odds ratio. There was a drop of ∼6% in external test set ROC-AUC for melanoma from an odds ratio of 1 in both training and test data to an odds ratio of 0.5/2 in the train/test data, respectively. Similarly, for chest x-rays, there was a drop of ∼7% in the external test set ROC-AUC for pleural effusion from an odds ratio of 1 in both training and test data to an odds ratio of 0.1/10 in the train/test data, respectively.

For melanoma prediction, we also observed that among the data sources that comprise the ISIC archive, the odds ratio for ‘female’ as a predictor for ‘melanoma’ varied from 0.475 to 1.185, confirming that reversals in the association between protected attributes and a prediction target indeed occur naturally in medical data. To further motivate this analysis, we also considered real-world melanoma classifiers, which are available for public use as smartphone apps. We analysed two classifiers, Scanoma and Smart Skin Cancer Detection (SSCD), to identify whether they relied on protected attributes for the melanoma prediction task. First, we first took the trained classifiers and tested them on a synthetic test set with a correlation between sex and melanoma with varying odds ratios. For both Scanoma and SSCD, we observed that the performance fluctuated as the strength of the odds ratio deviated from unity. Specifically, we saw a decreasing trend in performance as the odds ratio increased (ROC-AUC dropped by 8.73% for Scanoma and by 11.07% for SSCD, Figure 4). These declines indicated that both commercial classifiers also leveraged demographic encodings for disease predictions. To confirm this hypothesis, we then retrained the classifiers using an equalized training set with no correlation between sex and melanoma, essentially removing the dependence on demographic encodings. For both Scanoma and SSCD, we observed that performance remained consistent on the test set across varying odds ratios; in fact, performance *increased* compared to the earlier setting. We suspect that the retraining data for both classifiers came from a distribution that better matched the test set distribution than the original training data; however, the fact that the performance fluctuations were mitigated indicates that the original classifiers did, in fact, rely on demographic encodings that degraded their downstream performance for the clinically relevant classification tasks.

**Fig. 4.**
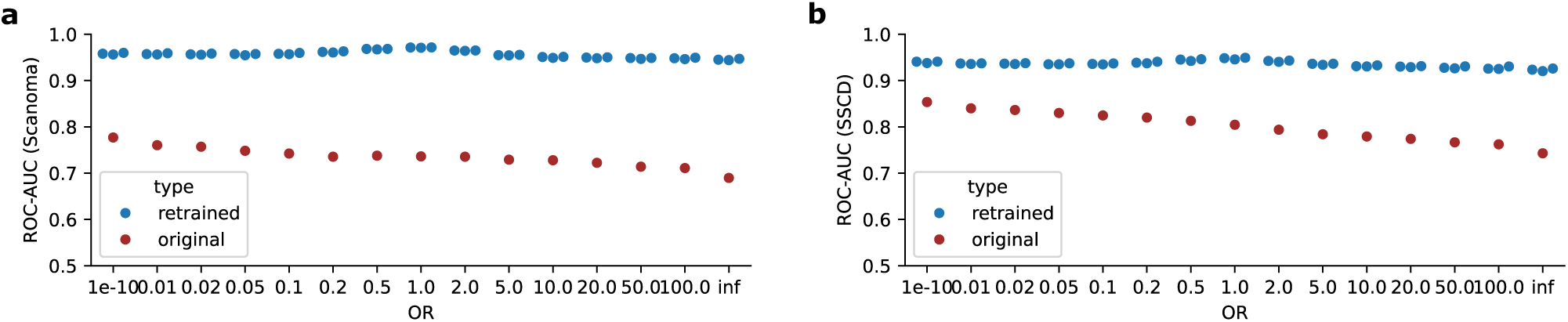
Analysis of the reliance on protected attributes for melanoma prediction for two real-world classifiers, Scanoma and SSCD. (**a**) Performance of Scanoma on the melanoma prediction task using a synthetic test set with varying strengths of association between sex and melanoma based on the odds ratio. The performance fluctuated as the odds ratio deviated from unity. On retraining Scanoma with an equalized training set without a correlation between sex and melanoma, the performance improved drastically. (**b**) Performance of SSCD on the melanoma prediction task. Similar trends were observed for this classifier as well.

### Exploration of statistical associations with the protected attributes

As an initial exploration toward understanding how the classifiers could identify protected attributes, we examined statistical associations with the available metadata characteristics, measured using the odds ratio for predicting female dermoscopic lesions and black chest x-rays in the training and testing sets. We found a few characteristics that associate dermoscopic lesions with sex and chest x-rays with race, albeit weakly.

For dermoscopic lesions, multiple diagnoses were weakly associated with sex. Some of these associations persisted from the training data, where an association must be present for the classifier to learn it, to the test data, where the association must persist to benefit performance. These include an association between female sex and solar lentigo, dermatofibromas or nevi, and an association between male sex and seborrheic keratoses (Supplementary Table 1). Since prior studies have successfully identified diagnoses from dermoscopic images,^20^ a classifier could, in principle, then leverage this knowledge to help identify a patient’s sex. Considering the dermoscopy method used for image acquisition, there was a weak association between non-contact polarized images and male sex (Supplementary Table 3). For chest x-rays, a similar trend was observed, with a few diagnoses weakly associated with race. These included an association between the black race and cardiomegaly or pneumonia, and an association between the white race and fracture or pneumothorax (Supplementary Table 2). Overall, however, associations between diagnoses or image acquistion techniques and the protected attributes appeared unlikely to account for the classifier’s performance on external test data considering that many associations were weak and that some associations were reversed between training and external test data.

In contrast, we observed a more consistent association of the protected attribute with patient age. For dermoscopic lesions, in the training data, patients aged 20-60 were enriched for females, while patients aged 5-15 and 65-85 were enriched for males (Figure 5b). In the external test set, patients aged 60-85 were also enriched for males, suggesting that a correlation between older ages and patient sex may persist across data sources. For chest x-rays, in both the training and the test data, patients aged 20-60 were enriched for the white race while patients aged 60-90 were enriched for the black race (Figure 5d), also showing a consistent correlation between the different ages and patient race which generalized across different hospital sites.

**Fig. 5.**
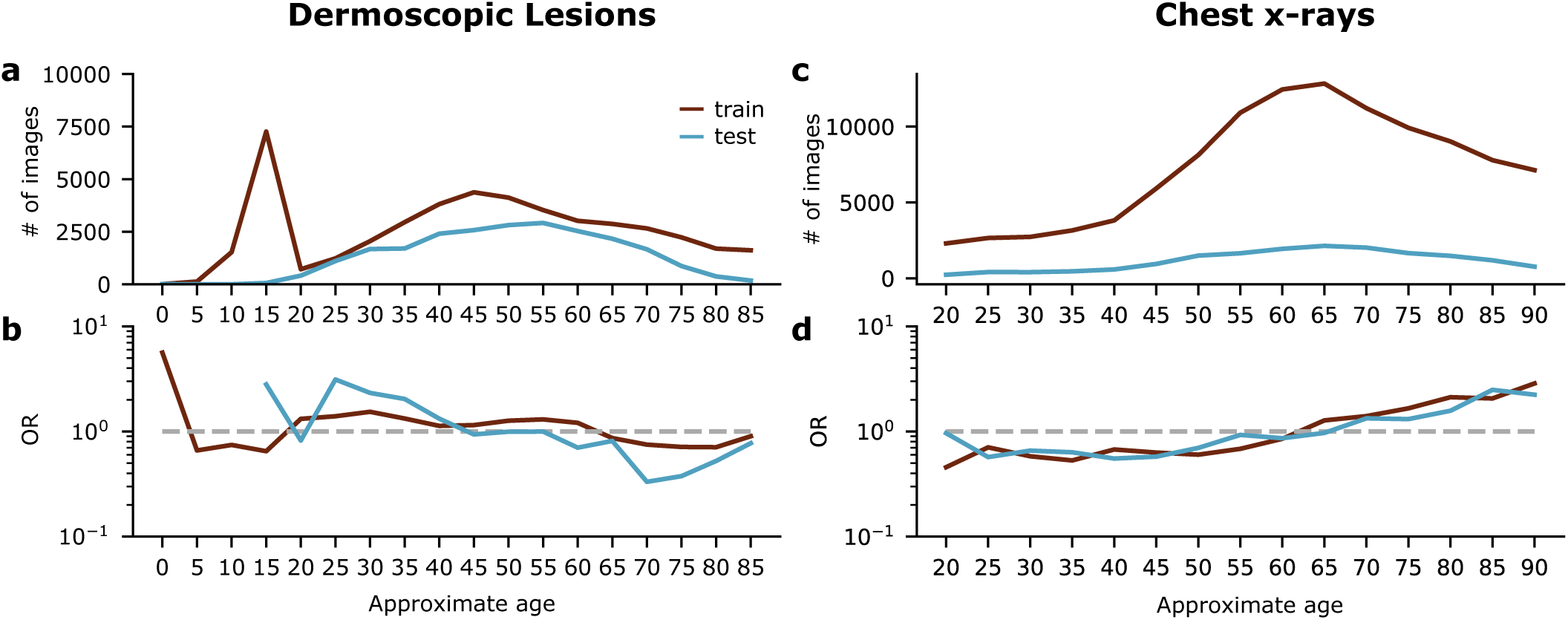
Association of patients’ sex and race with their age. **(a)** Histogram of patients’ ages in the training and test dermoscopic data. **(b)** Odds ratio (OR) for prediction of female sex on the basis of a patient’s age. **(c)** Histogram of patients’ ages in the training and test chest x-ray data. **(d)** Odds ratio (OR) for prediction of the black race on the basis of a patient’s age.

### Clustering-based analysis helps identify global artifacts

Using clustering analysis (see Methods), we examined visually similar clusters of images in the training data that differed the most in terms of the ratio of the protected attribute classes as predicted by the trained classifier.

For dermoscopic images, we considered the ratio of the predicted females to predicted males. Hair images differed strikingly between the two clusters, being highly prevalent in the cluster with more predicted males (Figure 6a). Since hair was a strong signal, to identify other signals, we equalized the training set by sub-sampling images so there was no statistical correlation between hair and sex. To label the images (that lacked detailed annotation), we manually annotated 500 female and 500 male images for the presence of hair and applied these hand-labeled images to train a classifier (ViT-Base architecture) for this task, achieving ROC-AUC of 0.96 on a held-out test set (90-10 train-set split). We then used that classifier to label the rest of the dataset. After sub-sampling, the new training set contained 11190 images without hair and 9230 images with hair for each of the female and male sexes, resulting in an odds ratio of 1. After retraining the sex classifier using the equalized training set, we performed the clustering analysis again; this time, stickers (i.e., small adhesive markers placed on the skin to indicate the location of lesions or areas of interest to guide biopsies, surgical excisions or other treatments) were identified as being more prevalent in the cluster with the highest number of predicted males, indicating that stickers could also be a potential signal associated with males by the sex classifier (Figure 6b). The exact type and color of the stickers could vary by hospital site, making them an easy ‘shortcut’^18^ to learn for predicting protected attributes (like sex).

**Fig. 6.**
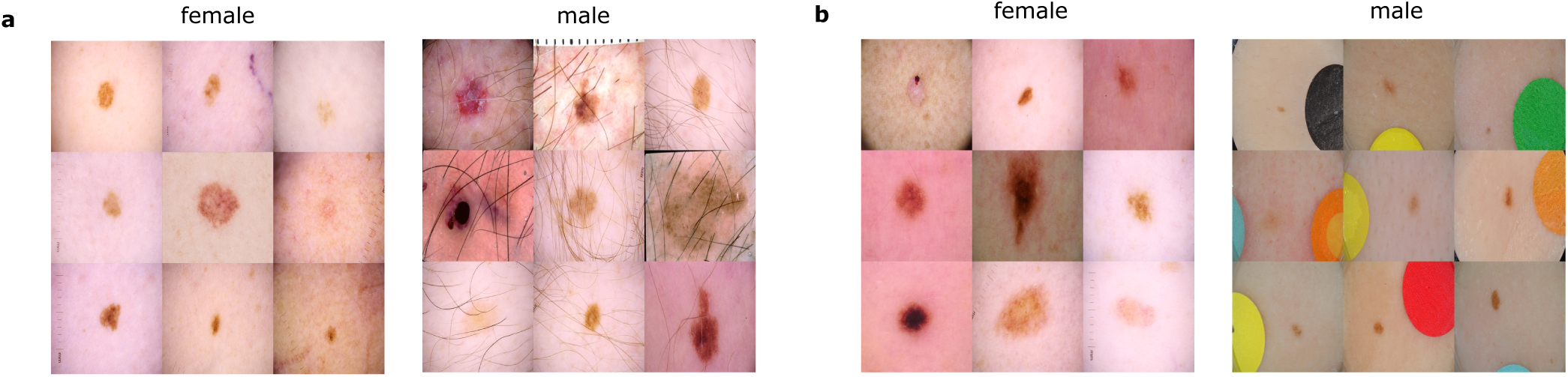
Clustering analysis of dermoscopic images. A sample of images from the visually similar clusters with the lowest and highest predicted male to female ratios for **(a)** the unequalized sex classifier, in which hair is identified as a potential signal, and **(b)** the sex classifier equalized for hair, in which the sticker is identified as a potential signal.

For chest x-rays, we considered the ratio of the predicted black race to predicted white race. The view position of the chest x-rays was strikingly different between the two clusters, with the cluster having more predicted black race being prevalent in x-rays with a lateral view position (Figure 7). Similar to the clustering analysis for dermoscopic lesions, we performed an additional equalization step to remove the correlation with the view position, however, it did not reveal any signals that stood out as being visually different between the new clusters.

**Fig. 7.**
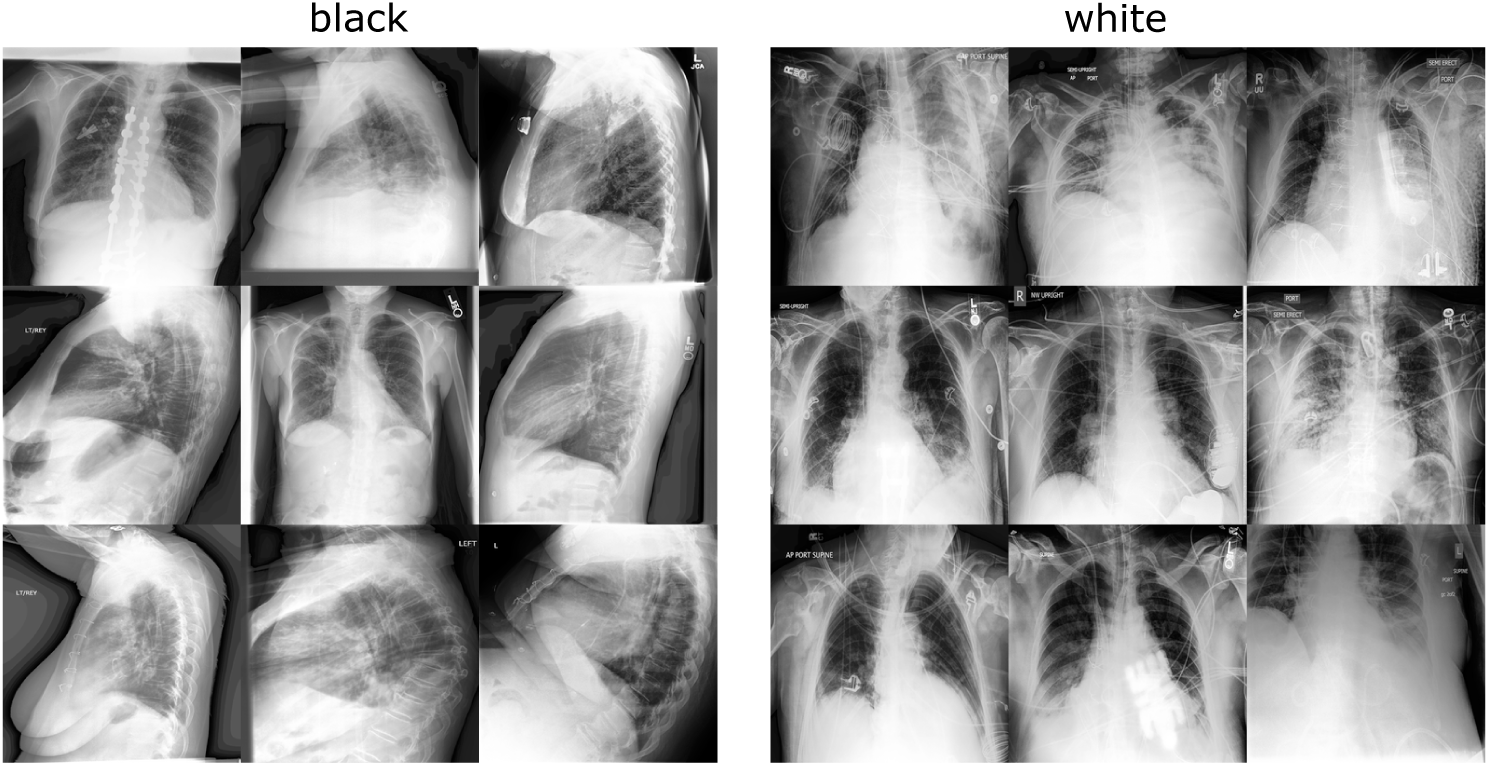
Clustering analysis of chest x-rays. A sample of images from the visually similar clusters with the lowest and highest predicted white to black ratios. The view position (frontal or lateral) was identified as a potential signal.

### Generative image AI reveals local prediction mechanism

Our generative techniques (see Methods) were able to produce realistic counterfactual images of dermoscopic lesions and chest x-rays. The distribution of images produced by the final network differed from the distribution of training images by a Fréchet Inception Distance (FID score)^21^ of 6.29 for the latent space optimization technique and 10.32 for the Explanation by Progressive Exaggeration (EBPE) technique for the dermatology data. For the radiology data, the FID scores were 35.24 for the latent space optimization technique and 44.37 for the EBPE technique. Some samples of the generated counterfactuals are shown in Figure 8. We generated a total of 200 pairs of counterfactuals from both the generative techniques that elicited a desired protected attribute prediction from the classifier, that is, a pair classified as either ‘female’ or ‘male’ and ‘black’ or ‘white’. The image pairs were then manually analysed by experts to identify signals in addition to those that were identified by clustering that could potentially be used by the classifiers for their predictions.

**Fig. 8.**
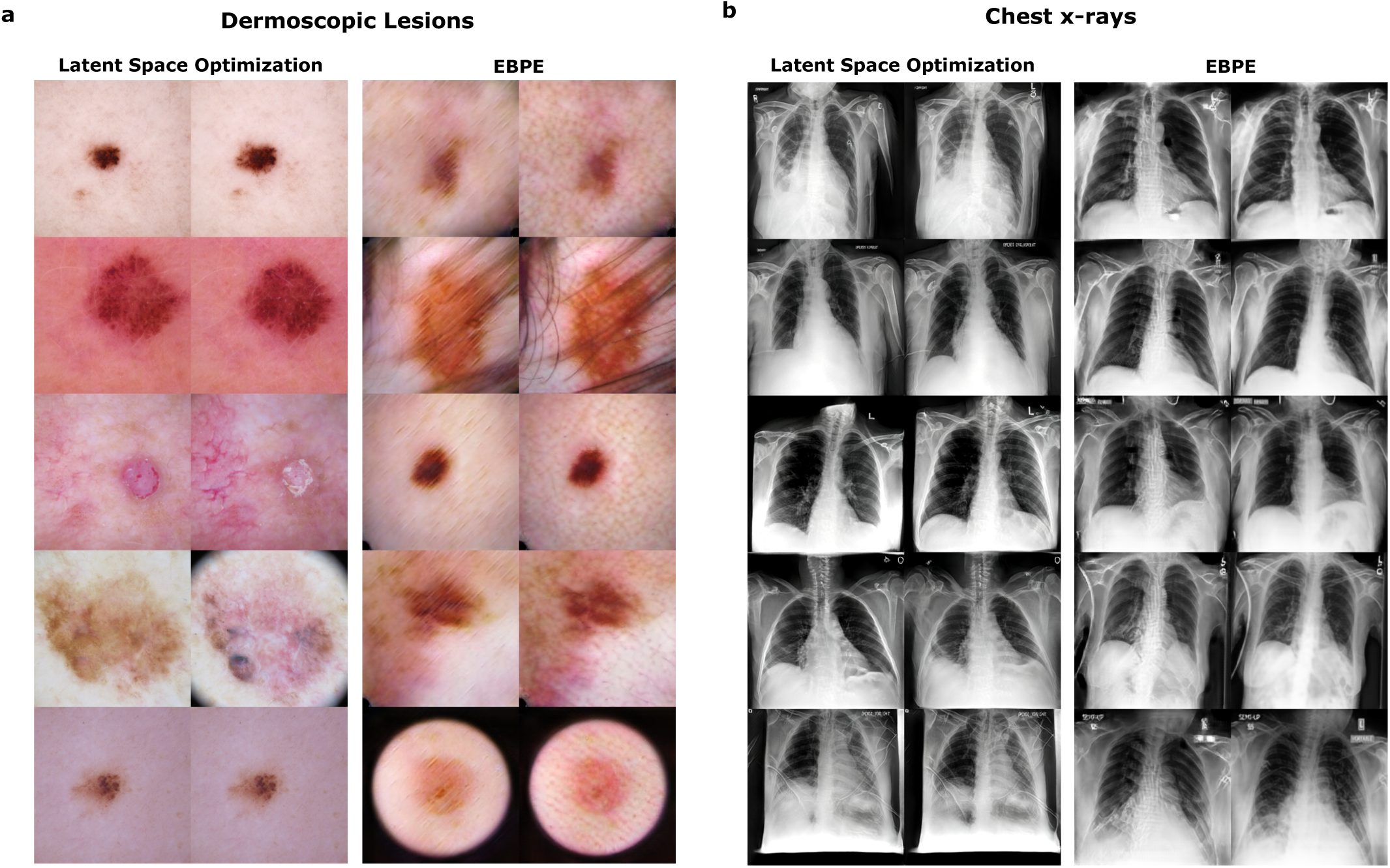
Examples of counterfactual images produced by our generative techniques. **(a)** For dermoscopic lesions, in each pair of images, the left one is the female counterfactual, and the right the male counterfactual. **(b)** For chest x-rays, in each pair of images, the left one is the black counterfactual and the right one is the white counterfactual.

Multiple visual signals in addition to the ones observed from the clustering analyses were identified for both dermoscopic lesions and chest x-rays (Tables 1 and 2) provide description of each signal, along with the method used to identify it and the direction of the protected attribute association). These signals were identified in at least 10% of the image pairs (for each generative technique) in the same direction (always from male to female or black to white), indicating possible correlations with the protected attribute. The dermoscopic lesion counterfactuals were analysed by two senior medical students (each of them analysed 200 pairs) while the chest x-ray counterfactuals were analysed by one radiology resident. To streamline the process of labeling images, we created a web app that displayed the counterfactual pairs and asked the experts to identify relevant signals (see Methods). Since we hypothesized that signals important for the classifier would be present in multiple counterfactual pairs, we discarded any signals that appeared in less than 10% of the pairs as being insignificant for the sex prediction task. Qualitative visualizations of the different signals are shown in Figure 9 for dermoscopic lesions and in Figure 10 for chest x-rays.

**Fig. 9.**
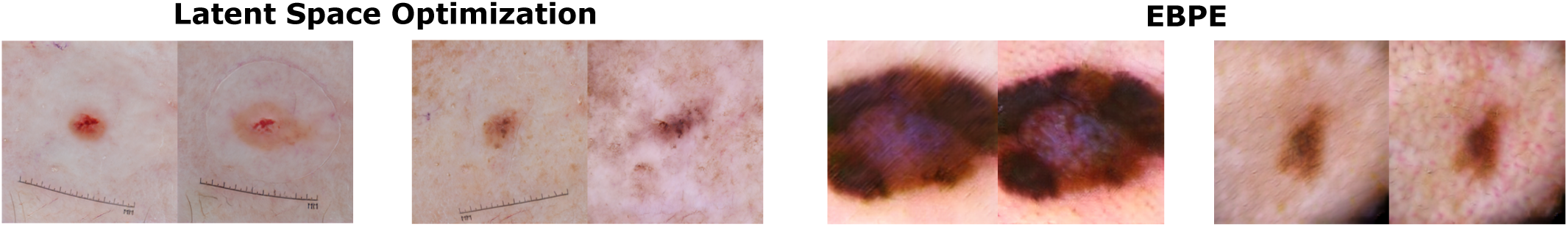
Signals identified from producing counterfactual pairs of dermoscopic lesions using the generative techniques. The two leftmost signal pairs are identified by the latent space optimization technique, and the two rightmost by EBPE. For each pair, the left lesion is the female counterfactual, and the right is the male.

**Fig. 10.**
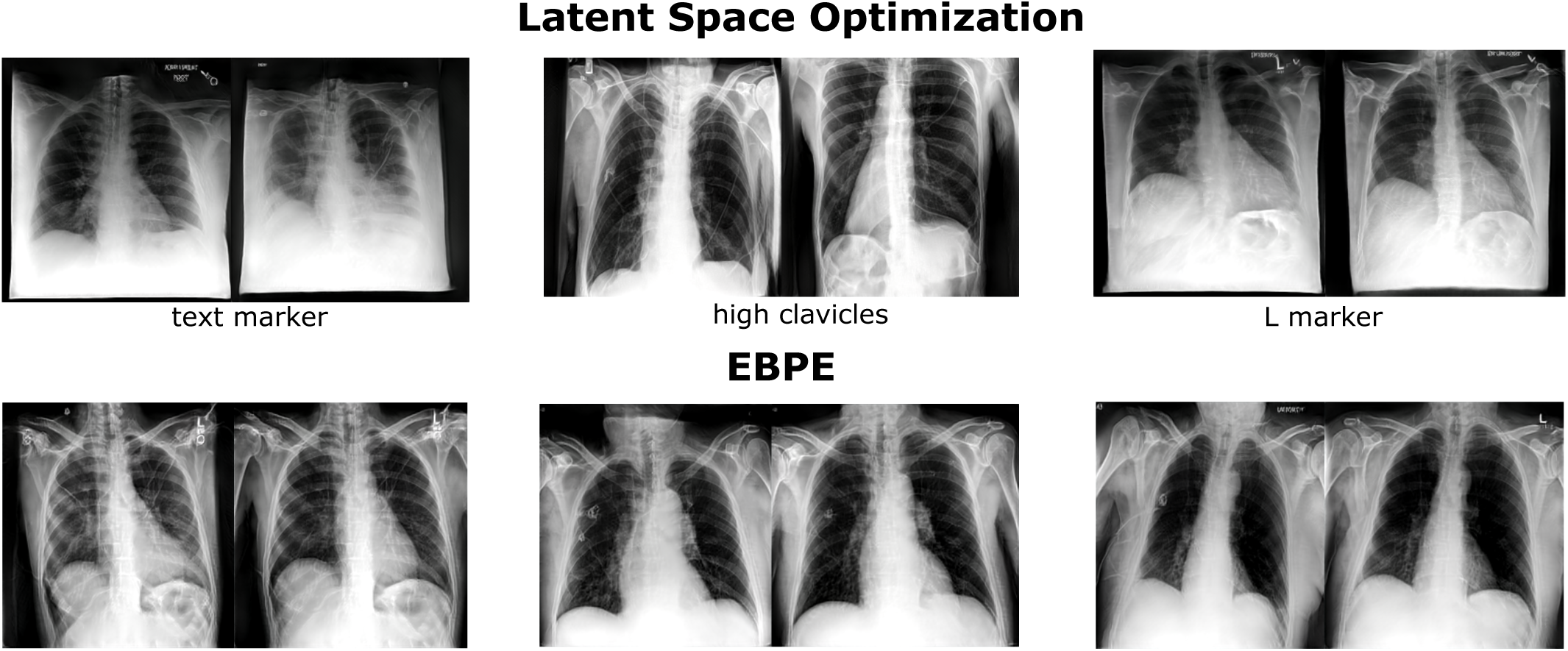
Signals identified from producing counterfactual pairs of chest x-rays using the generative techniques. The pairs in the top row are identified by the latent space optimization technique, and the ones in the bottom row by EBPE. For each pair, the left lesion is the black counterfactual, and the right is the white. The red markings highlight the differences based on the labeled signals. Most of the signals identified are either image artifacts like the markers and ecg leads or related to image acquisition parameters like patient positioning which can affect lung width and height.

**Table 1.** AI-specific signals for dermatology identified by analyzing the female and male counterfactual images obtained from the generative techniques. The description specifies what the signal corresponds to visually and the association indicates the sex in which the signal was more prevalent.

| Signal Identified | Description | Generative Technique | Association |
| --- | --- | --- | --- |
| dermoscopic gel | Visible dermoscopic gel, also known as immersion fluid, in the image used to enhance visualization of the skin lesion. | Latent Space Optimization | Male |
| ruler | The dermoscopic image has visible ruler markings that serve as a scale reference. | Latent Space Optimization | Female |
| pigmentation | Presence of color or pigment within the lesion, ranging from amelanotic to black in color. Darker pigmentation was found to be associated with males. | EBPE | Male |
| redness | The skin lesion or the background shows signs of reddish dots, indicating inflamed skin or erythema. | EBPE | Male |

**Table 2.** AI-specific signals for radiology identified by analyzing the black and white counterfactual images obtained from the generative techniques. The description specifies what the signal corresponds to visually and the association indicates the race in which the signal was more prevalent.

| Signal Identified | Description | Generative Technique | Association |
| --- | --- | --- | --- |
| L Marker | The L-laterality marker on a chest x-ray is used to indicate the left side of the patient's body. | Latent Space Optimization | Black |
| High Clavicles | The shoulders/clavicles are close to the top of the chest x-ray or out of the field of view | Latent Space Optimization | White |
| Text Markers | Annotations added to the chest x-ray that provide contextual information for record keeping, usually present at the top of the x-ray. | Latent Space Optimization | Black |
| Width | Mean of the widths of the left and the right lungs (mean calculation not needed for lateral x-rays) | EBPE | White |
| Height | Mean of the heights of the left and the right lungs (mean calculation not needed for lateral x-rays) | EBPE | White |
| ECG Lead | Electrocardiogram (ECG) monitoring electrodes or wires that are visible in the chest x-ray | EBPE | Black |

### ‘Removal via balancing’ successfully quantifies classifier performance

After applying a range of techniques, including statistical association analysis, clustering analyses, and generative modeling to identify putatively meaningful signals, we confirmed and quantified their importance using *removal via balancing* (see Methods). This technique quantifies the importance of a putatively important *query signal* for a classifier’s predictions in a particular test set. For example, how much does hair contribute to the prediction of sex from dermoscopic lesions? To implement this technique, we compared the model’s performance in the original test data to its performance in an alternate version of that test data in which the query signal was statistically independent of the prediction target. In other words, under the hypothesis that a classifier depended on a particular query signal, we would expect the classifier’s performance to decline when there was no difference in the query signal between target classes (female and male or black and white), and the degree by which the performance declines quantified the importance of that signal.

The removal via balancing technique required a numerical representation for each signal in each image; since these annotations were absent from the original data, we produced annotations using a hybrid approach, similar to the one used to equalize hair in the clustering analysis by training labeling classifiers (see Methods).

For the dermoscopic lesions, after quantifying the importance of the signals we previously identified (age, hair, sticker, redness, ruler, pigmentation, and gel), we found that they collectively explained about 42% of the classifier’s performance; the largest single contributor was hair, which alone explained about 37% of the classifier’s performance. Other attributes, e.g., age, redness, and presence of rulers, explained a smaller but non-negligible proportion of the classifier’s performance (Figure 11a, Table 3). Attributes like pigmentation, sticker, and gel explained minimal performance, suggesting that these attributes were either not leveraged by the classifier (false positives) or they were not prevalent in the test set. For example, the test set contained only two images with stickers, indicating that the classifier could not rely on this signal for prediction in the test set despite learning the signal during training. Some of the identified signals, such as hair, can be explained by physiological sex-based differences since males typically have more body hair than females. However, the other identified signals, e.g., age, redness, and ruler, do not conform to known biological insights and can be specific to the training data or image acquisition parameters.

**Fig. 11.**
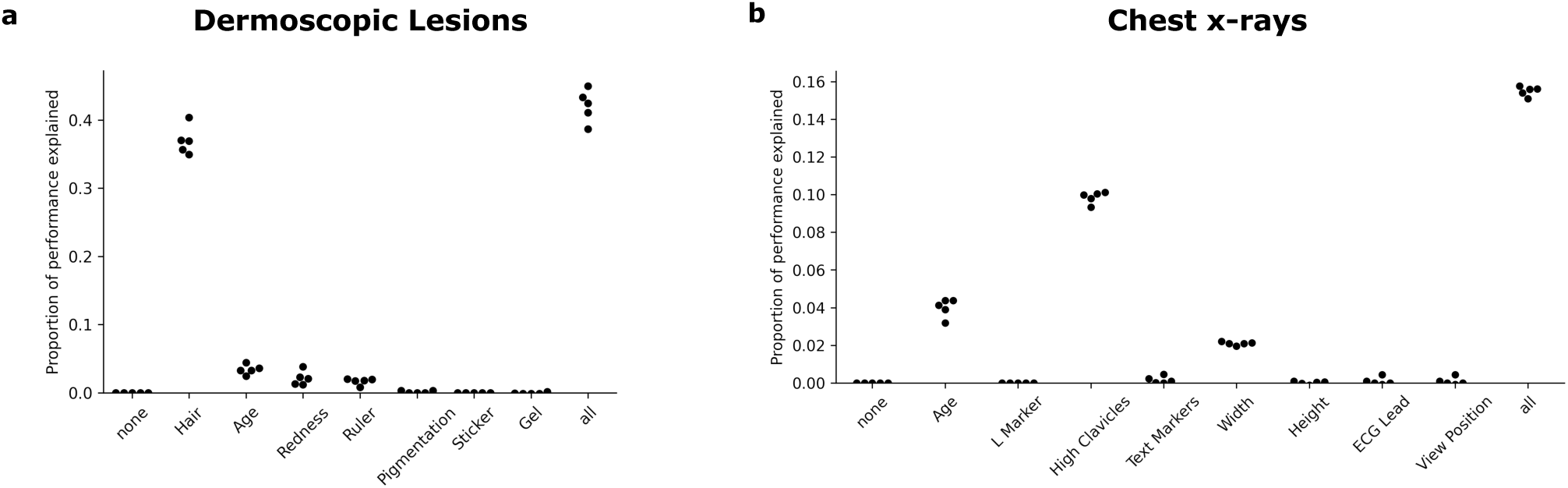
The proportion of performance explained by signals identified in our prior experiments. The proportion of ROC-AUC explained across five replicates for **(a)** Dermoscopic Lesions and **(b)** Chest x-rays. **Note:** The scale of the y-axis is different on the two plots for easier visualization of small quantification values; the total proportion of performance explained to lower on chest x-rays compared to dermoscopic lesions.

**Table 3.** Proportion of the ROC-AUC explained along with standard deviations across five replicates by the signals identified from the XAI analysis which have non-zero quantification for both the dermatology and radiology modalities.

| Dermoscopic Lesions |  | Chest x-rays |  |
| --- | --- | --- | --- |
| Signal Identified | Quantification | Signal Identified | Quantification |
| Hair | $36.983 \pm 1.64$ | Age | $3.995 \pm 0.61$ |
| Age | $3.405 \pm 0.57$ | High Clavicles | $9.855 \pm 0.38$ |
| Redness | $2.133 \pm 0.82$ | Width | $2.092 \pm 0.12$ |
| Ruler | $1.633 \pm 0.39$ | **All** | **$15.398 \pm 0.32$** |
| **All** | **$42.093 \pm 1.87$** |  |  |

For the chest x-rays, after similarly quantifying the importance of all the signals identified (age, view position, L-marker, high clavicles, text markers, width, height, ECG lead), we found that they collectively explained ∼15% of the the classifier’s performance with the position of the clavicles accounting for the largest contribution (∼10%). The age of the patient and lung width also contributed to small proportions of the performance. Similar to dermoscopic lesions, many of the identified attributes did not contribute much to the classifier’s performance. For example, the test set only contained ∼600 (out of ∼26,000) lateral-view images, making any signal related to the view position that may be learned by the classifier during training ineffective on the test set. This further highlights the importance of quantifying the signals identified since there can false positives or distribution shifts.

## Discussion

As AI technologies are increasingly becoming part of the clinical workflow, it becomes vital to ensure that these models work as designed and do not encode sensitive information that can be leveraged to cause discriminatory performance in downstream tasks, an unacceptable vulnerability. An important step towards mitigating this behavior is establishing a generalizable framework for understanding the extent to which this issue is prevalent and why it occurs. In this work, we propose such a framework, called DREAM, and apply it to two medical modalities, dermatology and radiology.

As the first step of our DREAM framework, we demonstrated that we can achieve unexpectedly high performance in predicting sex from dermoscopic images and race from chest x-rays, which aligns with prior work^8,9^. We then showed how these high-performing predictions can be detrimental to the prediction of clinically relevant downstream tasks by disease classifiers^22,23^. Furthermore, our analysis of commercial melanoma prediction models revealed that real-world systems already display this vulnerability, with marked performance drops under manipulated protected attribute correlations. To shed light on the exact mechanisms these protected attribute classifiers use to achieve such high performance, the second step of DREAM involved a preliminary statistical analysis, which revealed a correlation between age and both patient sex and race. In addition, by combining methods like clustering and counterfactual image generation with expert analysis, we were able to identify visual signals for both the modalities that the classifiers could potentially leverage to make protected attribute predictions. As the final step, we used our novel ‘removal via balancing’ technique to quantify each of the signals identified.

Our findings indicate that some of the predictive signals models exploit reflect physiological differences (e.g., hair presence in males lesions compared to female lesions). Other signals reflect image acquisition artifacts or dataset-specific biases that persist across populations or settings. Notably, certain signals — such as age, ruler markings, clavicle position, patient positioning (which can impact lung width and height), and laterality markers — are unlikely to have direct biological relevance and may serve as shortcuts that generalize across hospital sites and the models opportunistically exploit. This has also been shown in prior work, with the acquisition parameters of chest x-rays influencing the prediction of race by AI classifiers^14,15^. Since some of the identified signals (like stickers, markers, etc.) could be manually manipulated in the images, we further verified their impact and associations with the protected attributes by artificially introducing them or removing them from the images and measuring the change in the classifier prediction (see Supplementary Methods).

The fact that only part of the model performance (∼42% for dermoscopic lesions and ∼15% for chest x-rays) could be explained by these signals also suggests that there may exist residual non-visual signals, i.e., subtle image patterns imperceptible to human observers but learnable by AI classifiers. Since our analysis relies on visual scrutiny, it may miss such non-visual, AI specific signals. The existence of such features underscores the difficulty of this task and highlights the need for greater scrutiny and transparency in model development and deployment. As an initial step toward detecting such non-visual signals, we performed an experiment using frequency spectrum analysis on the dermoscopic lesion images and were able to empirically confirm the existence of such features (see Supplementary Methods).

Our results also generalized across model families: further analysis of classifiers trained with transfer learning (Supplementary Section 4.1) and with dermatology foundation model embeddings exhibited similar behaviors and relied on similar signals to predict protected attributes (Supplementary Section 4.2). This indicates that demographic encoding is not the result of a specific architecture or training paradigm, but rather a broader property of data-driven feature learning in high-dimensional visual domains.

It is worth noting that we used GANs for generating images and although diffusion models^24,25^ have been shown to generate more realistic images, they have not had as much success in counterfactual generation. Prior work that attempted to use diffusion models for this task^26^ focused on natural images, not medical ones, which are arguably more difficult to generate. Furthermore, diffusion model training has more computational overhead and the sampling is more expensive compared to GANs since they require multiple iterations of the reverse process to create one sample. GANs have been successfully used for counterfactual generation of medical images^4,5,15^, and the quality of the generated images for both modalities (in terms of the FID score) is satisfactory for our analysis.

In sum, this work motivates and introduces a comprehensive interpretability framework called DREAM to identify and quantify visual signals, by leveraging expert analysis, in medical images that lead to the prediction of protected attributes. Although this study focused on two modalities, DREAM is flexible and applicable to other modalities (e.g., retinal fundus images, pathology slides), which we anticipate could reveal similar vulnerabilities. As AI becomes increasingly embedded in clinical decision-making processes, ensuring generalization and fairness must be considered an integral part of the development process.

## Methods

### Data preparation

To study how AI classifiers predicted protected demographics from medical images, we focused on the prediction of a patient’s sex from a dermoscopic image of their skin and the prediction of a patient’s race from their chest x-ray.

Dermoscopic images offer a magnified view of the skin and typically lack anatomical landmarks (eyes, nose, fingers, etc.) that could offer a route to the identification of a patient’s sex (male or female). We leveraged data contained in the ISIC archive^27–30^, specifically the 2019 and 2020 ISIC challenge datasets, which consists primarily of dermoscopic images collected by global medical professionals along with associated metadata on diagnoses, demographic characteristics, and details on image acquisition. For our study, we excluded non-dermoscopic images. We then partitioned that data based on provenance as encoded by the ‘attribution’ metadata label, intended to credit images under Creative Commons Attribution licenses but often also used to provide information on the image acquisition site (Supplementary Table 4). This partitioning scheme minimized the chance of overlap in patients between the training and test data; it additionally provided a more robust test scenario for domain shift analysis since spurious associations in training data were unlikely to persist in the different hospitals and geographic regions of the test data. After preprocessing and partitioning, the train set had 45924 images and the test set had 23461 images.

Chest x-rays are images generated of the chest to create a picture of the structures and organs in the chest, including the lungs, heart, and rib cage. They also lack any features that may indicate race, like skin color. As our training set, we used the CheXpert dataset^16^, which contains chest x-rays collected from the Stanford Health Care center. As our testing set, we use the MIMIC-CXR dataset^17^, which contains chest x-rays collected from a different hospital site, the Beth Israel Deaconess Medical Center in Boston. We filtered the images to those containing demographic information like patient race and age and considered the black and white races for our analysis to be consistent with a binary prediction task. After filtering, the train set had 109962 images and the test set had 37272 images.

### Model training

As the first step of DREAM, we trained vision transformer-based models (ViT-Base architectures)^31^ for both the prediction tasks. To do so, we started with classifiers pre-trained on ImageNet^32^ and then replaced the 1000-class linear classification head with a new linear head suited for binary prediction. For training, we held out 10% of the training data (selected at random) as a validation set. We optimized the network using an Adam optimizer with learning rate 10^−5^, *β*_1_ = 0.9, and *β*_2_ = 0.999, with a mini-batch size of 512 and cross-entropy loss as our optimization criterion. We optimized the models for 30 epochs, reducing the learning rate by a factor of 0.2 (i.e., *lr_new_* = 0.2×*lr_old_*) if the model’s loss did not improve for 5 epochs. Finally, we used the epoch with the highest ROC-AUC in the validation data for all subsequent experiments. We repeated this procedure for 5 replicates.

### Clustering based analysis

The second step of DREAM involved leveraging multiple explainable AI methods to identify signals in the dataset that the classifier relies on to predict sex. For the clustering based analysis, we grouped images in the train dataset based on their visual similarity using the K-means clustering algorithm^33^ implemented in the scikit-learn Python package (K=20). To obtain clustering features, we used the first 50 principal components derived by running Principal Component Analysis (PCA) on the embeddings of the penultimate layer of a Resnet50 model^34^ (pretrained on ImageNet^35^), which is a layer capable of capturing the visual and structural similarities between different images.

Once we had the clusters, we calculated the ratio of the protected attribute labels (predicted males to predicted females for dermoscopic images and predicted white to predicted black for chest x-rays) for each cluster and picked out the two with maximum and minimum ratios corresponding to the male-dominant and female-dominant clusters respectively for dermatology, and the white-dominant and black-dominant clusters respectively for radiology. Then, clinical experts analyzed a subset of 100 images selected uniformly at random from the two clusters to identify signals that differ. Visually clustering the images before comparison allowed us to hypothesize that the signals identified would be significant to the model prediction and not just identified by chance.

### Generation of counterfactuals

Another XAI tool we used to investigate the signals that potentially guide the prediction of AI classifiers is called *counterfactual image generation*.^36–38^ Counterfactual images are synthetic images that reveal the basis of an AI classifier’s decisions by altering attributes of a reference image to create a similar image that prompts a different prediction of a protected attribute from the classifier. For example, consider a pair of generated counterfactual images from a real dermoscopic lesion, one that is predicted by the AI classifier as belonging to a female and the other that differs in some visual signals and is predicted by the classifier as belonging to a male. Then, assuming that we ensure all differences between the counterfactuals push the AI classifier’s predictions in the desired direction (female to male), we may infer that the classifier uses those signals as part of its reasoning process for predicting sex from dermoscopic lesions.

We wanted to create pairs of counterfactual images that (1) appeared “realistic” in the sense that they lie on the manifold of training images, (2) produced the desired target prediction from a classifier, such as a prediction on the opposite side of the decision threshold as the original image, and (3) were similar to the original image in the sense that the original image can be approximately reconstructed by passing an altered, generated image back through the generator. To generate counterfactual images, we employed two different methods to get a diverse range of counterfactuals and obtain a wide spread of potentially important signals for the sex and race prediction tasks, as follows.

### Latent space optimization

In the first method, we followed prior work^39–41^ and simply optimized the image using gradient descent to elicit a desired response from the classifier. To ensure the image remained realistic, we optimized the latent representation of that image in the latent space of a generative adversarial network rather than doing so directly in pixel-space. (Figure 12) Since we were interested in broadly understanding the predictions of the classifier rather than explaining the classifier’s predictions for a specific output, we utilized randomly generated images as our references, eliminating the need for an encoder network as was used in prior efforts^39^.

**Fig. 12.**
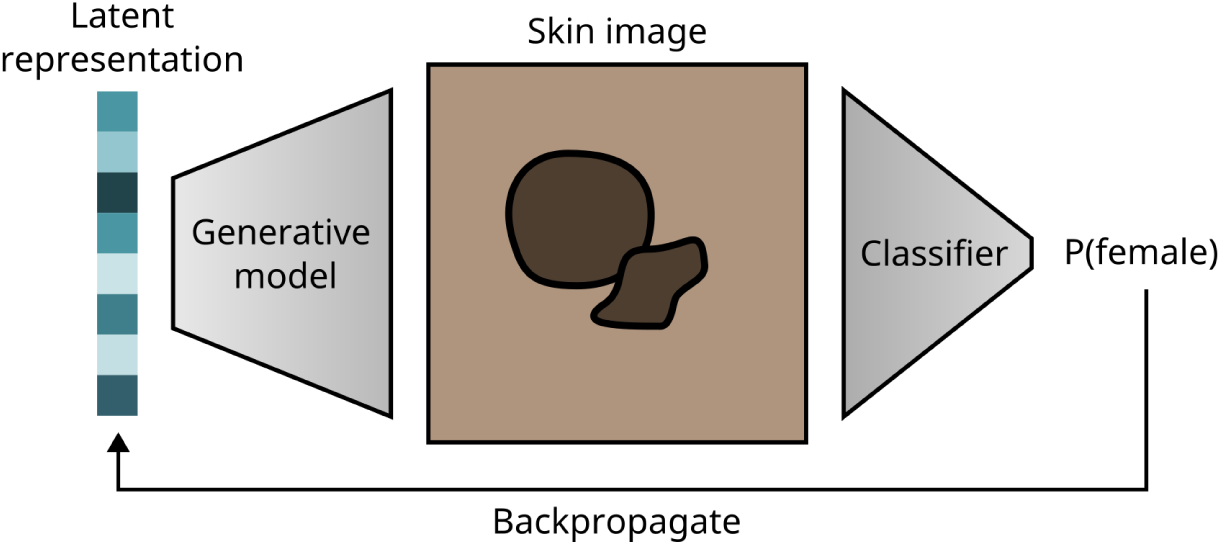
Latent space optimization for generating counterfactuals using a StyleGAN. Given a StyleGAN trained on either dermoscopic images or chest x-rays and a classifier trained to predict the corresponding protected attribute, we started with a random latent vector in the GAN’s latent space. We then used the generator and the classifier’s prediction to optimize the latent representation to generate an image that elicited the desired output probability from the classifier. Throughout the training process, only the latent representation was updated while keeping both the generator and the classifier frozen. Dermoscopic images are shown here for illustration purposes.

We generated a pair of counterfactual images by first choosing a random latent vector 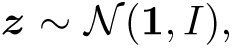 where **1** is a *d*-dimensional vector of 1s and *I* is the *d* × *d* identity matrix. Given a generator 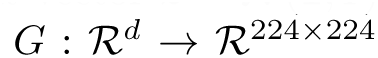 and classifier 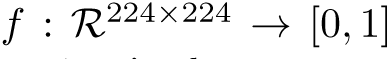 that quantified the probability of the image representing a female patient’s lesion or a black patient’s chest x-ray, we performed gradient descent on *z* to optimize *f* (*G*(*z*)). Based on any given *z*, we generated a female counterfactual lesion or a black counterfactual chest x-ray by minimizing −*f* (*G*(*z*)) until *f* (*G*(*z*^′^)) *>* 0.95 (where *z*^′^ represents the updated latent vector) and a male counterfactual lesion or a white counterfactual chest x-ray by minimizing *f* (*G*(*z*)) until *f* (*G*(*z*^′^)) *<* 0.05. Since the optimization is deterministic and the optimization problem is not convex, a portion (∼30%) of initial vectors *z* failed to produce a counterfactual based on the preceding criteria; we stopped optimization after a maximum of 500 steps and excluded these from further analysis.

During optimization, we use a learning rate of 0.05 (after hypterparameter tuning) for both the modalities. The generator and the classifier were kept frozen during the optimization procedure, and only the latent representation was updated. For the generator *G*, we chose a styleGAN2 (a generative adversarial network, or GAN) with adaptive discriminator augmentation.^42^ More details about the training procedure are provided in Supplementary methods.

### Explanation by Progressive Exaggeration

In the second, more rigorous method, we generated counterfactual images using a variant of the technique *Explanation by Progressive Exaggeration*(EBPE),^43^ as described in prior work.^5^ These updates seek to improve image quality, stabilize training, and restrict the altered attributes to those that would drive a classifier toward a different prediction (e.g., avoiding any adversarial perturbations or altering confounding attributes).

Formally, let X ⊂ [0, 1]*^d^*^2^ represent a set of images drawn from some data manifold 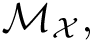 is the horizontal and vertical resolution of the (square) images, and let *f*: [0, 1]*^d^*^2^ → [0, 1] be a classifier that outputs the probability that a given image belongs to a female patient for dermoscopic lesions and a black patient for chest x-rays. Our goal was to obtain a generator 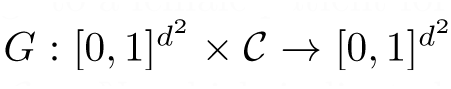 that produced a counterfactual image 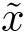 when given an input image *x* and a condition *c* ∈ C ⊂ N, which indicated the target output probability that the classifier should produce when evaluated on the counterfactual image 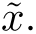.

As in the original implementation of EBPE, our condition *c* was a discrete value that indexed a “bin” in the discretized output space of the classifier *f*; we chose C = {0, 1*,…,* 9} with corresponding target outputs in the bins {[0, 0.1), [0.1, 0.2)*,…,* [0.9, 1]}. The requirements listed above then meant that (1) the range of the generator *G*(X, C) was contained in the data manifold M_X_, (2) the classifier’s prediction for the generated image *f* (*G*(*x, c*)) was approximately equal to the target output (in our case, the bin’s center at *c/*10 + 0.05), and (3) if *f* (*x*) fell within the bin indexed by *c*, then *G*(*G*(*x, c*^′^)*, c*) ≈ *x* for each *c*^′^ ∈ C.

To obtain a generator with these properties, the generator *G* was optimized in conjunction with a discriminator network *D*: [0, 1]*^d^*^2^ → R, which attempted to distinguish real from generated images. We updated the discriminator so that it did not depend on a condition *c*. The original discriminator implementation attempted to differentiate generated from real images that elicited a particular prediction from the classifier, which could encourage generated images to appear similar to the subset of real images that potentially include changes that do not alter the output of the classifier. In contrast, the modified discriminator implementation instead attempted to differentiate generated images from any real image such that it only encouraged the generated images to appear similar to real images.

To reflect this update, the following functions were chosen for the loss of the discriminator *L_D_*and the generator *L_G_*. In the following equations, the random variables *X* and *C* take values in X and C and are distributed uniformly over X and C; *θ_D_* and *θ_G_* are the parameters of the discriminator and generator, respectively; *b*: [0, 1] → C returns the bin index *b*(*f* (*X*)) of the output of the classifier;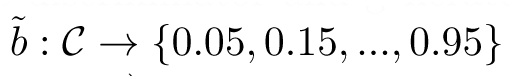 returns the center of the bin at index *C*; and *D_KL_* is the Kullback–Leibler divergence (Figure 13).

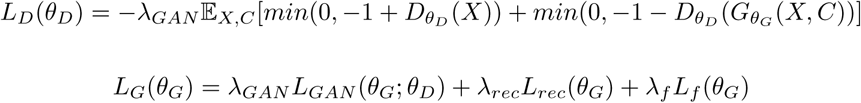

**Fig. 13.**
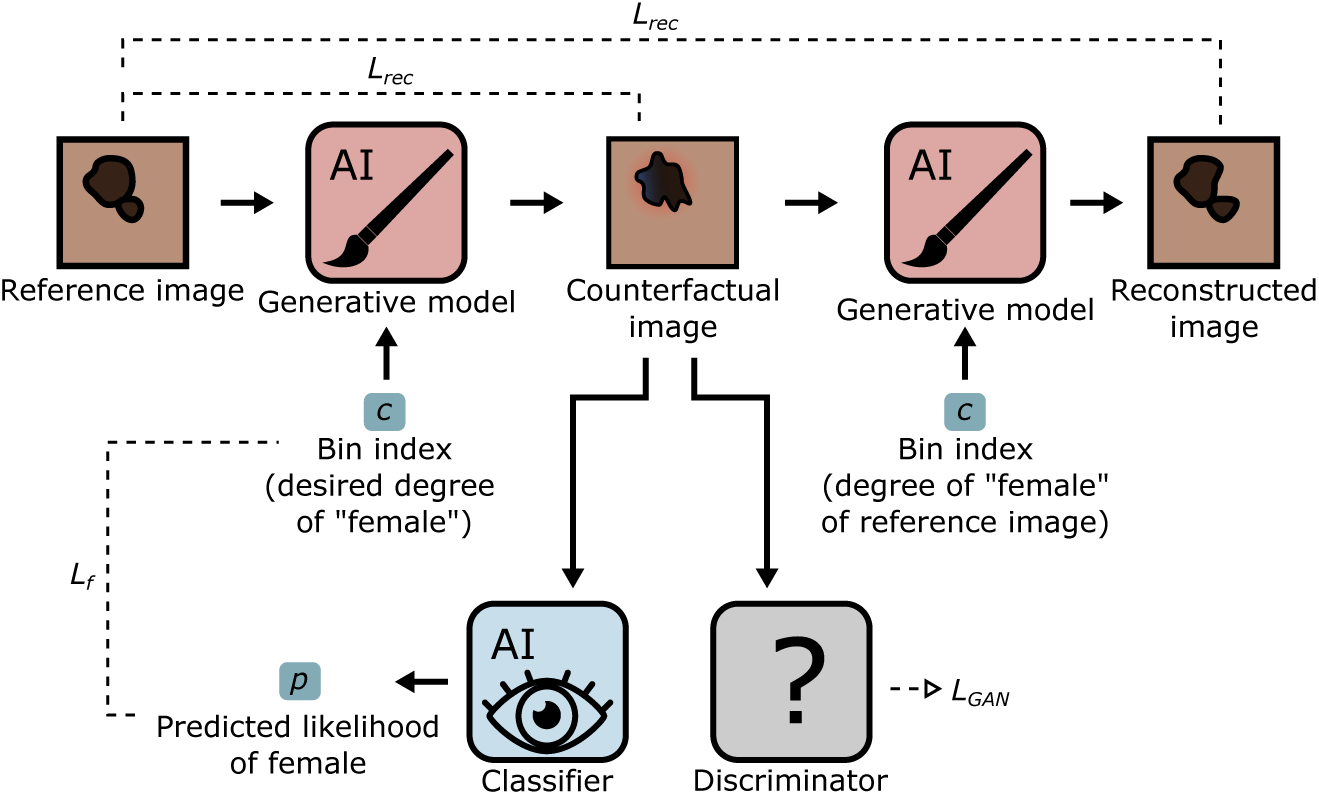
The GAN setup used during training with the modified EBPE technique. Given a reference image and a bin index representing the desired prediction from the classifier, the generative model created a coun-terfactual image. The loss term *L_f_* enforced that the counterfactual, when evaluated by the classifier (which was frozen), generated the corresponding output as specified by the bin index. The counterfactual was also passed to the discriminator, which attempted to discern whether it represented a real or generated image and thus enforced realism of the counterfactuals (via *L_GAN_*). The reconstruction L1 loss *L_rec_*enforced that the counterfactual was similar to the reference image. It consisted of two steps: (1) The counterfactual was passed back to the generative model, along with the bin index that corresponded to the classifier’s prediction on the reference image, in an attempt to reconstruct the reference image (*L_rec_*, top). (2) We also attempted to reconstruct the reference image with only a single pass through the generator (*L_rec_*, lower) by again passing a bin index that matched the classifier’s output on the reference image. Dermoscopic images are shown here for illustration purposes.

The individual components of *L_G_* are:

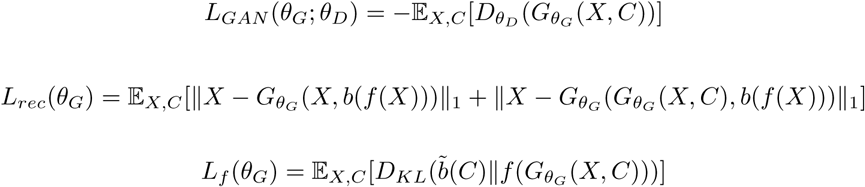

We chose the extreme values for *c* (*c* = 0 and *c* = 9, corresponding to classifier outputs of 0.05 and 0.95 respectively) in order to generate counterfactuals that were predicted to strongly elicit the desired protected attribute predictions. In addition to the introduction of a non-conditional discriminator, *G* was also updated to use an architecture similar to that used in CycleGANs^44^ to improve image quality. The optimization procedure and further training details are described in Supplementary Methods.

### Expert evaluation of counterfactuals

The next step in DREAM was to identify specific visual image signals that the classifiers possibly used to predict protected attributes. For this, we asked two senior medical students to analyze dermoscopic counterfactual images and a radiology resident to analyze chest x-ray counterfactual images generated by the two preceding methods and determine which aspects of each image were altered, implying that they could be potential signals that affected the classifier’s decisions.

To help the experts evaluate the counterfactuals, we developed a graphical interface in the form of a web app hosted on Amazon Web Services (AWS). Each web app page displayed a single pair of counterfactual images, with both having opposite predictions from the classifier for the protected attribute of interest. The counterfactual pair to be displayed was generated by either the EBPE technique or by the latent space optimization technique (selected at random), and the expert evaluators were not told which technique was used.

The experts first analyzed the pair of counterfactuals and then answered questions, including (1) Did the images look realistic? (2) How did the two counterfactuals differ from each other in terms of visual attributes? The evaluators could enter a free text response for the second question. They could also save their progress and return to the app to pick up where they left off. The app also provided a slider which the evaluators could use to linearly interpolate from one counterfactual to the other for easier examination of the subtle changes that occurred between the two. Figure 14) shows a visualization of the interface and the list of questions that were asked.

**Fig. 14.**
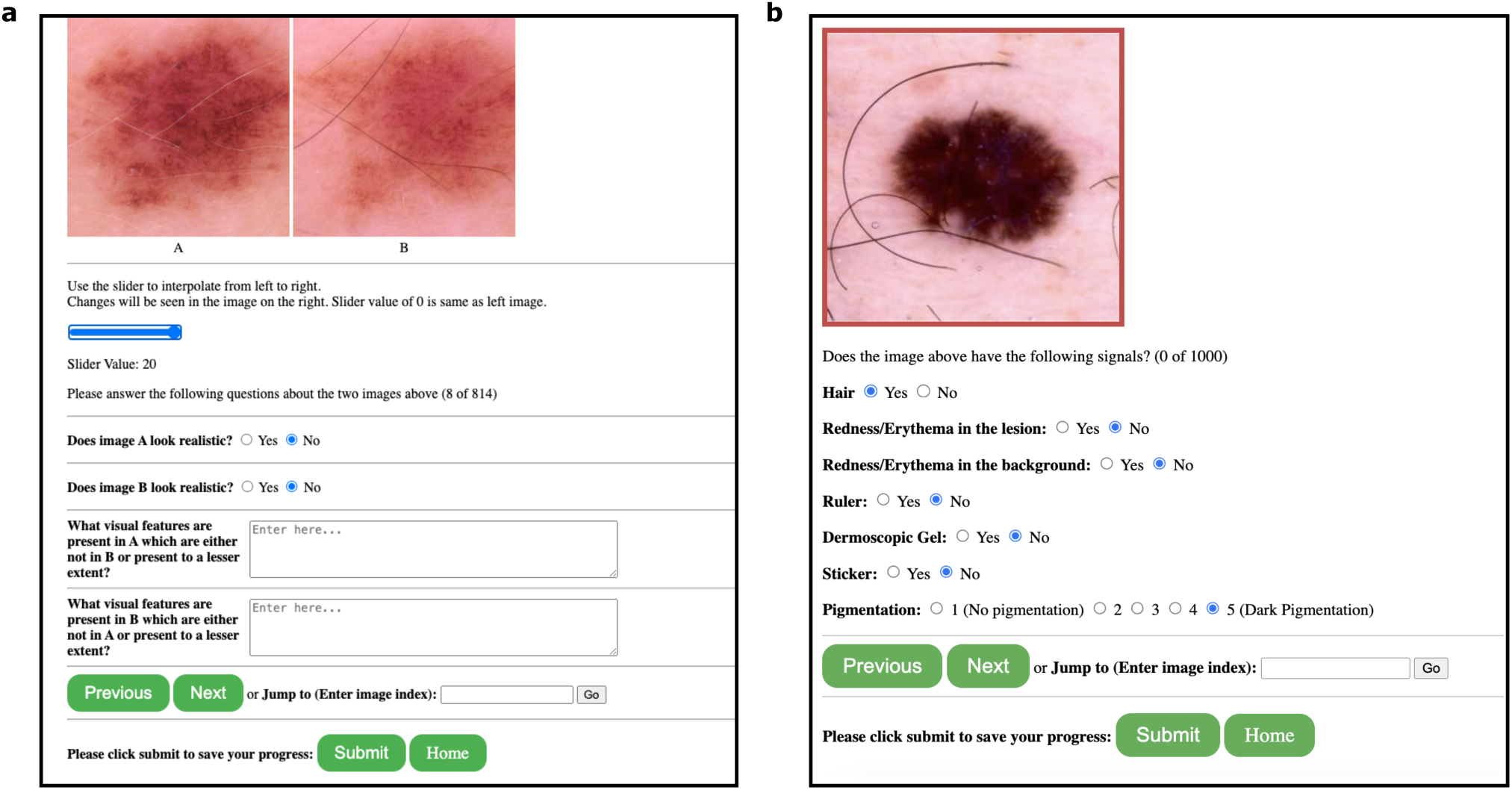
Web apps used to identify signals from the counterfactual images and label those signals in the testing set for quantification. **(a)** Screenshot of the UI used to identify signals that were visually different between a pair of counterfactuals. The slider could be used to interpolate between the counterfactuals to assist in highlighting the differences. **(b)** Screenshot of the UI used to label the identified signals. The user labeled the presence or absence of each signal. For pigmentation, we also let the user select from a scale from no pigmentation to dark pigmentation. The web-app for dermoscopic images is shown here for illustration purposes.

After we analyzed all counterfactual pairs, we discarded the pairs for which either of the images looked unrealistic and grouped similar visual features that were recorded in multiple images in the same direction.

### Quantification of explained performance with ‘removal via balancing’

Once we identified the signals that could potentially exist, the final step of DREAM involved quantifying the amount of the AI classifier’s performance that could be explained by a putatively important signal (or signals), which we term the ‘query signal(s)’, defined as the drop in predictive performance when the signal(s) was ‘removed’ from the test data, in the sense that the query signal was balanced with respect to the prediction target. To measure this drop, we developed a new method called *removal via balancing.* Specifically, to ‘remove’ a query signal *A*, we updated the test data to form a new pseudo-population in which *A* ⊥ *Y*, where *Y* is the protected attribute being predicted (for example, a patient’s sex). Our scheme required that each signal be represented by a scalar or vector in R*^n^* (where *n* ∈ N), e.g., as a scalar quantification of that signal or a one-hot encoded vector. Our goal was to weight each sample by the reciprocal of its propensity, 1*/P* (*Y* |*A*), which, in alignment with prior work^45–48^ on inverse probability treatment weighting (IPTW) in the field of treatment-effect estimation, provided a pseudo-population with the desired property that *A* ⊥ *Y*. Since the true propensity was not known, we followed prior work^47^ and estimated it via a logistic regression 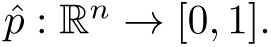 We then assigned the weight 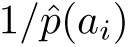 to sample *i* with vector of signals *a_i_*(Figure 15).

**Fig. 15.**
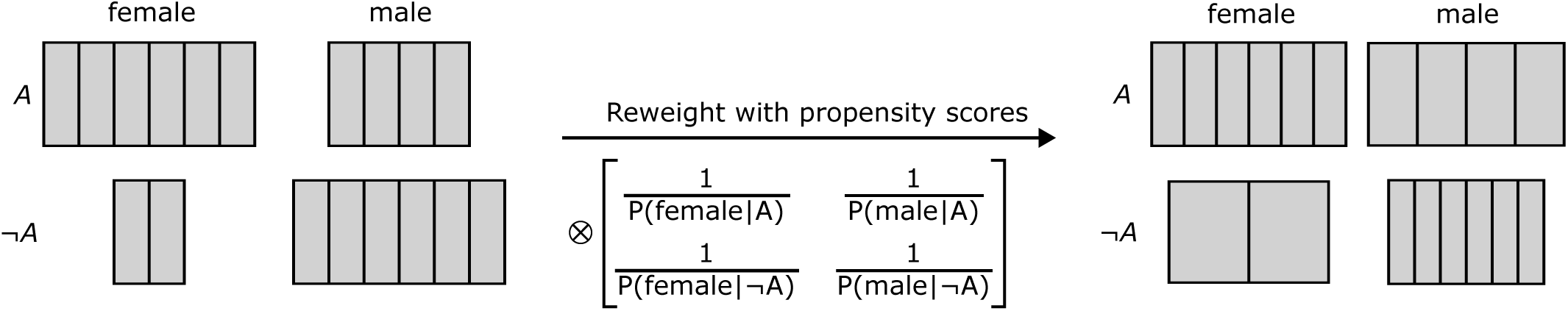
Visual representation of the quantification technique. Given a query signal with a strong association with the one of the targets, we estimated it’s propensity for each class by using a logistic regression. Then, we weighed each of the samples by the inverse of its propensity score, creating a pseudo-population with the desired property of removing the association between the query signal and the target class. The illustration is shown for sex prediction from dermoscopic lesions.

We caution that despite borrowing balancing scores from the causal inference literature, our technique does not aim to infer causal relationships between query attributes and the model’s predictions. In our view, no direct analogy can be drawn between our use of balancing scores and their use in treatment effect estimation. Importantly, when our technique removes a query signal via balancing, correlated signals may also be (partially) removed. For instance, if two signals correlate perfectly in the test data, our technique cannot differentiate which signal is important for a classifier. In this way, our technique defines signals on the basis of how they appear in the test data; e.g., in the extreme case, two semantically different but perfectly correlated signals are effectively defined as a single signal for the purposes of our analysis.

We quantified the *proportion of performance explained* as the ratio of the performance decline after balancing to the maximum possible performance decline (to random performance of ROC-AUC=0.5):

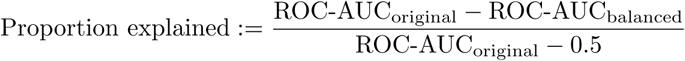

This method needed signal labels for all the images in the test data to train the logistic regression model, which were not available in the metadata. To generate these labels, we created another web app hosted on AWS. This app showed an image from the test set and asked the expert evaluators to label it with the identified signals (Figure 14b). We did this for a subset of the images (500 for each category of both sex and race) and trained a classifier (ViT-Base architecture, ROC-AUC: ∼0.90) to label the remaining test data. For the pigmentation signal identified in the dermatology counterfactual analysis, we asked the evaluators to pick from a scale of 1 to 5, with 1 indicating no pigmentation and 5 indicating very dark pigmentation. To binarize the signal, all values above 3 were considered to be darkly pigmented and those below to have light pigmentation.

For radiology, one of the signals identified from the counterfactual analysis was the dimension of the lungs (width and height). To obtain these values for the test set, we trained a segmentation model to segment the lungs from the chest x-rays. Specifically, we used a pre-trained segmentation model called Ternausnet^49^ which is a U-net^50^ with a VGG11^51^ encoder. We used the model to segment lung regions from input images and generate binary masks. For each mask, the two largest contours (corresponding to the two lungs) were found using OpenCV^52^, and the chest x-ray image was divided vertically to distinguish between left and right lung regions based on the centroid of each contour. A bounding box was then drawn around each contour and the width and height of these bounding boxes were recorded as the dimensions for each lung. For lateral chest x-rays, only one contour was obtained.

## Data availability

Images used in this study were obtained from publicly available repositories. ISIC images are available at https://challenge.isic-archive.com/data/. We used the images from the 2019-2020 ISIC challenges since they had protected attribute information in the metadata. The CheXpert Chest X-ray dataset is available at https:// stanfordaimi.azurewebsites.net/datasets/8cbd9ed4-2eb9-4565-affc-111cf4f7ebe2 and the MIMIC-CXR dataset is available at https://physionet.org/content/mimic-cxr-jpg/2.1.0/.

## Code availability

Our code, implemented in PyTorch, is available at https://github.com/suinleelab/protected-attrib-prediction. It includes the dataset preprocessing pipeline, detailed step-by-step documentation for running our DREAM frame-work, and a demo of the web-app that was used to identify the signals from counterfactual pairs.

## Data Availability

Images used in this study were obtained from publicly available repositories. ISIC images are available at https:
//challenge.isic-archive.com/data/. We used the images from the 2019-2020 ISIC challenges since they had protected attribute information in the metadata. The CheXpert Chest X-ray dataset is available at https://stanfordaimi.azurewebsites.net/datasets/8cbd9ed4-2eb9-4565-affc-111cf4f7ebe2 and the MIMIC-CXR dataset is available at https://physionet.org/content/mimic-cxr-jpg/2.1.0/.

https://challenge.isic-archive.com/data/

https://stanfordaimi.azurewebsites.net/datasets/8cbd9ed4-2eb9-4565-affc-111cf4f7ebe2

https://physionet.org/content/mimic-cxr-jpg/2.1.0/

## Supplementary Methods

### 1 Additional experiments to verify identified signals

To further validate the identified signals in addition to the quantification, we manually manipulated the signals to add or remove them from the images and tested their effect on the predictions.

### 1.1 Dermoscopic Images

For dermoscopic lesion images, we focused on the “redness” and “stickers” signals which were easier to edit in the images. The other signals were more diffused and, therefore, difficult to edit in the images by direct manipulation.

#### 1.1.1 Effect of redness

The presence of red dots on the dermoscopic images was identified as one of the potential signals associated with the male sex by the trained classifier. To replicate the effect of having red dots, we manually added red dots to the existing images from the test set. We first started with a blank image of the same width and height as the lesion and added 100 red dots of a fixed radius to random locations within the image. Then, we blended the original dermoscopic image with it to simulate the presence of red dots (Supplementary Figure 1a). We then observed the change in prediction accuracy and the male predicted probability before and after adding the red dots for the dermoscopic lesions in the male and female subgroups.

Considering the subgroup of ground truth male lesions, both the accuracy and the male predicted probability increased after adding the red dots (Supplementary Figure 1b). The accuracy increased 21% from ∼0.718 to ∼0.871 while the male predicted probability increased 20% from ∼0.716 to ∼0.861. Considering the subgroup of ground truth female lesions, the accuracy decreased and the male predicted probability increased after adding the red dots (Supplementary Figure 1c). The accuracy decreased 54% from ∼0.702 to ∼0.325 while the male predicted probability increased from 111% ∼0.312 to ∼0.659. Both of these results indicated that images with redness were associated with male lesions by the classifier and further validated this signal identified through counterfactual image analysis.

#### 1.1.2 Effect of stickers

From the clustering analysis, stickers were identified as a potential signal that the classifier could use to predict sex from dermoscopic lesions, with male being associated with the presence of stickers. Since stickers are artifacts which are not part of the skin lesion, they are easier to manually manipulate. We used different techniques to add and remove stickers from the images in the dataset.

To remove the stickers from the images which had stickers, we first had to segment them within the image. We used the recently developed Segment Anything Model (SAM)^53^ to segment out the stickers in the images and it worked well for our purposes (accuracy of 90% from 500 manually tested images). Once we had the masks for the stickers, we inpainted them with the color of the skin to emulate the removal of stickers. However, the skin had different coloration in different regions, so using a simple mean color of the unmasked pixels did not look realistic. Instead, to do the inpainting, for each mask, we considered the 5 closest pixels that were bordering the sticker mask and took the mean color of those to inpaint the masked region (Supplementary Figure 2a). To emulate the addition of stickers to the images without stickers, we selected a set of three stickers from existing dermoscopic images with each sticker representing different sticker patterns that were observed in the datasets. We then overlaid one of them selected at random on each of the images either at the top, bottom, left, or right sides (Supplementary Figure 2b). Once we had a set of images in which the stickers were either inpainted or overlaid, we observed the change in the accuracy and the male predicted probability of the classifier before and after the manipulation for the male and female subgroups.

Considering the case in which we started with dermoscopic images which had stickers and then inpainted them using the technique mentioned above, for the subgroup of male lesions, the accuracy decreased 15% from ∼0.903 to ∼0.770 and the male predicted probability also decreased 14% from ∼0.867 to ∼0.749. For the subgroup of female lesions, both of accuracy and the predicted probability remained almost the same (Supplementary Figure 2c). This indicated that removing stickers from images makes them more ”female-like” to the classifier, which corroborated the findings from the clustering analysis.

In the case where we started with dermoscopic images which didn’t have stickers and manually added stickers to them, for the subgroup with male lesions, both the accuracy and male predicted probability remained the same before and after the addition. For the subgroup with female lesions, the accuracy decreased 27% from ∼0.860 to ∼0.624 and the male predicted probability increased 206% from ∼0.153 to ∼0.468. This further confirmed the observation that adding stickers to images made them appear more “male-like” to the classifier.

### 1.2 Chest X-Rays

For chest x-rays, we focused on the ‘High Clavicles’ and ‘L Laterality Markers’ since those are also signals which were easier to edit in images.

#### 1.2.1 Effect of High Clavicles

From the latent space optimization technique, high clavicles were identified as a potential signal and they were associated with white chest x-rays. To replicate the effect of high clavicles, we started with all the chest x-rays that were labeled as not having high clavicles. Then we cropped the top 50 pixels of the image and resized it to the size of the original image. This would make it seem the the clavicles have been shifted up (Supplementary Figure 3a). We then observed the change in prediction accuracy and the predicted probability for the black race before and after the manipulations for each of the white and black subgroups.

Considering the subgroup of black chest x-rays, both the accuracy and the predicted probability decreased after the clavicles were shifted higher (Supplementary Figure 3b). The accuracy decreased 16% from ∼0.797 to ∼0.667 and the predicted probability decreased 20% from ∼ 0.770 to ∼0.617. This result indicated that the chest x-rays with high clavicles are associated with the white race, which aligned with what was revealed during the counterfactual evaluation by experts. Considering the subgroup of white chest x-rays, there was no significant change in both the accuracy and the predicted probability before and after the manipulation (Supplementary Figure 3c).

#### 1.2.2 Effect of L-Laterality Markers

From the latent space optimization technique, the L-Laterality markers were also observed as being a potentially important signal associated with the black race, although they did not contribute much to the performance when assessed by the quantification technique. To replicate the effect of having L-Laterality markers, we first cropped out icons for the different styles of the markers that were present in the testing set. Then, we selected the chest x-rays which were labeled as not having those markers by our classifier and manually pasted the icons on top of those images. We selected a style at random for each image and also randomly placed it either on the top-left or the top-right side of the image since those were the locations where any laterality markers were annotated (Supplementary Figure 4a). Finally, we observed the change in prediction accuracy and the predicted probability for the black race before and after the manipulations for each of the white and black subgroups.

Considering the subgroup of black chest x-rays, there was no significant change in the accuracy and the predicted probability before and after the markers were added. Considering the subgroup of white chest x-rays, there was a slight decrease in accuracy and the predicted probabilities. The accuracy decreased 3% from ∼0.946 to ∼0.916 and the predicted probability increased 28% from ∼0.085 to ∼0.119. These results also aligned with the observations from the counterfactual analysis and the small change indicated that the signal might not be leveraged strongly by the classifier for the race prediction task.

## 2 Training details for the counterfactual generation methods

Here we provide additional training details on the two methods we employed to generate counterfactuals, latent space optimization and explanation by progressive exaggeration (EBPE).

### 2.1 Latent Space Optimization

To train the styleGAN2 that was used as the generator, we used all images from the ISIC 2019 and 2020 datasets, which were resized such that the short edge measured 256 pixels and then center-cropped to 256×256 pixels. We fine-tuned the model starting from a checkpoint pre-trained on Flickr Faces High Quality 256 (FFHQ256)^54^. During training, we augmented the training data by randomly, horizontally flipping images. We optimized the networks using the Adam optimizer with a learning rate of 0.0025, *β*_1_ = 0, *β*_2_ = 0.99, and batch size of 64. For adaptative discriminator augmentation, we set the target to 0.6.^55^ The training was done using a NVIDIA RTX 2080TI GPU with 11 GB memory.

### 2.2 Explanation by Progressive Exaggeration

To optimize the discriminator and the CycleGAN-based generator, we followed the PyTorch reference implementation^5^ and used an Adam optimizer with a learning rate of 2 × 10^−4^, *β*_1_ = 0, and *β*_2_ = 0.9, with a mini-batch size of 32. To prevent the discriminator from outpacing the generator, we trained the discriminator for 5 mini-batches for each mini-batch that the generator was trained, and we applied spectral normalization to the discriminator’s parameters. To avoid overfitting, we also applied data augmentation, including random cropping and random brightness modifications. To choose the hyperparameters *λ*, we followed the reference implementation and chose *λ_GAN_*= 1 and *λ_f_* = 1. To balance the magnitude of the generator’s alterations such that the counterfactuals were similar to original images but still contained perceptible differences (based on manual visual analysis of images), we chose *λ_rec_*= 3 after gradually relaxing the *λ_rec_* term from the value *λ_rec_* = 100 as suggested in the original publication. The generative models for each classifier and for each dataset were all trained using identical parameters. Comparison of counterfactuals generated by independent re-trainings of a generative model preserved which attributes varied between the male and female counterfactuals for dermatology or the black and white counterfactuals for radiology. The generative models were trained for 500 epochs on a NVIDIA RTX 2080TI GPU with 11 GB memory.

## 3 Frequency Spectrum Analysis

We were not able to explain 100% of the performance for either of the two modalities that were analysed since we were only able to identify visual signals that the classifiers leveraged. AI classifiers could also be sensitive to features that humans can’t directly visualize in the original images and they could use those differences to make predictions. To this end, we followed prior work^9^ and performed a frequency spectrum analysis on the dermoscopic images to break them into their component frequencies and evaluated the image frequencies for which the classifier had a higher sensitivity for the task of sex prediction. Spatial frequency describes how quickly pixel values change in space, with the higher frequency features representing sharp transitions between different regions, such as edges, lines, textures, and the lower frequency features representing gradual changes in color or intensity, such as those found in backgrounds or large, uniformly colored areas.

To obtain the component frequencies from an image, we first applied a fast Fourier transform (fft) to the image which converted it to the frequency spectrum. Then we shifted the zero-frequency component to the center of the spectrum to make filtering easier. In the shifted Fourier transform, the center of the image represented the highest frequencies and they decreased as we moved away from the center. Following this, to perform the frequency filtering, we created two circular masks, one with a radius of 5 pixels and the other with a radius of 50 pixels. For the smaller circular mask, we applied a high-pass filter, causing only the highest frequencies present inside the circle to pass through while filtering the rest out. For the bigger circular mask, we applied a low-pass filter, causing the highest frequencies within the circle to get filtered out and only allowing the lower frequencies outside the circle to pass through. After applying these filters, we then used the inverse fast Fourier tranform (ifft) to convert the images back from the frequency domain to the pixel domain (Supplementary Figure 5a). The high-pass filter resulted in images in which only high frequency features (like hair) were retained, while the low-pass filter resulted in images in which only low frequency features were retained, which created a softening or blurring effect.

Once we had the filtered images, we evaluated them using the trained classifier (Supplementary Figure 5b). Unsur-prisingly, the original images without any filtering performed the best. However, contrary to expectations, the images obtained from the high-pass filter (ROC-AUC of 0.615 ± 0.008, mean ± standard deviation) outperformed the images obtained from the low-pass filter (ROC-AUC of 0.523 ± 0.009, mean ± standard deviation) even though most of the image was not visible after the high-pass filtering. Low-pass filtering only caused the image to get blurred while retain-ing most of the structural and color-based variations, but this still caused the performance to drop significantly. On the other hand, we lost most of the visual information using the high-pass filter but it still retained enough information to have high performance. These results further validated that hair was an important signal that the classifier relied on to make predictions along with other frequency-based features which may not be visually identifiable to humans.

## 4 Generalization across model families

Here we show that other types of classifiers can also leverage visual signals similar to those identified for ViT-based classifiers to predict protected attributes with unexpectedly high performance.

### 4.1 Melanoma classifier embeddings encode demographic information

In a practical scenario, deep learning classifiers would not be trained specifically for the sex prediction task, but would rather be trained for disease prediction. However, even such classifier embeddings can encode some information for protected attribute classification. To test this out in the dermatology domain, we first trained a ViT-based classifier to predict melanoma versus melanoma look alikes. Then, we used transfer learning to retrain just the linear head to predict sex instead of melanoma, while keeping the rest of the model weights frozen (Supplementary Figure 6a). We used the same data splits as the ones used for training the ViT-based sex classifier. If the embeddings obtained from the melanoma classifier rely on demographic information, the transfer-learned classifier should also perform well on the sex classification task.

We observed that the transfer-learned classifier was able to predict sex with a high performance that was comparable to the performance using the original ViT-based classifier (ROC-AUC of 0.742 ± 0.015; mean ± standard deviation; Supplementary Figure 6b). Next, using the ‘removal via balancing’ technique, we quantified the same signals that were identified using the original ViT-based classifier. All the signals that were identified earlier were also important for the transfer-learned classifier, with hair explaining a bulk of the performance (about 45%). Overall, we were actually able to explain more of the performance of the transfer-learned classifier compared to the original classifier (56% versus 42%). This indicates that the transfer-learned classifier can encode demographic information to a larger extent, and it can be used for predicting clinically relevant downstream tasks like melanoma.

### 4.2 Foundation Models Encode Demographic Information

More recently, foundation models^56^ are gaining popularity over specialized models trained for specific downstream tasks due to their ability to generalize across a wide range of tasks, leveraging extensive pretraining on diverse, largescale datasets. One such family of foundation models is based on CLIP (Contrastive Language-Image Pre-Training)^57^, initially developed by OpenAI by training a self-supervised contrastive learning objective on a vast corpus of images sourced from the internet paired with textual descriptions to learn a joint embedding space. Such multi-modal models are especially effective in the clinical domain where there is a lack of structured, specialized datasets with ground truth. This has led to the development of numerous CLIP-like biomedical foundation models by finetuning using domain-specific datasets in the form of image caption pairs^58–64^.

These foundation models can also potentially encode biases similar to fully-supervised models and learn information about protected attributes like sex and race which can result in spurious correlations and be detrimental to downstream performance. To test this hypothesis, we considered one such dermatology foundation model called MONET^64^, trained on dermatological images paired with natural language descriptions obtained from PubMed articles and medical textbooks. We used the trained image encoder to get the image embeddings and fit a linear classifier on top for the sex prediction task. We used the same training, validation, and testing sets as the ones used for the ViT-based sex classifier.

The classifier had high performance on the external test set (ROC-AUC of 0.798 ± 0.002; mean ± standard deviation; Supplementary Figure 7a). Using the ‘removal via balancing’ technique to quantify the importance of the same signals that were identified with the ViT-based classifier, we observed that the same signals were also important for this classifier. ‘Hair’ again accounted for the largest contribution, explaining about 55% of the classifiers performance, which was significantly higher than the performance explained of the ViT-based classifier. ‘Age’ and ‘Redness’ accounted for a smaller but non-negligible proportion of the performance while ‘Ruler’ was not important for the classifiers prediction, contrary to what was observed with the ViT-based classifier. Overall, we were able to explain about 65% of the classifier’s performance, which is significantly higher than what we observed with the ViT-base classifier (Supplementary Figure 7b). This experiment shows that even foundation models do encode demographic information and their learning mechanisms seem to be similar to those of fully-supervised classifiers, with similar signals being important for prediction albeit with varying quantification.

---

**Supplementary Fig. 1.**
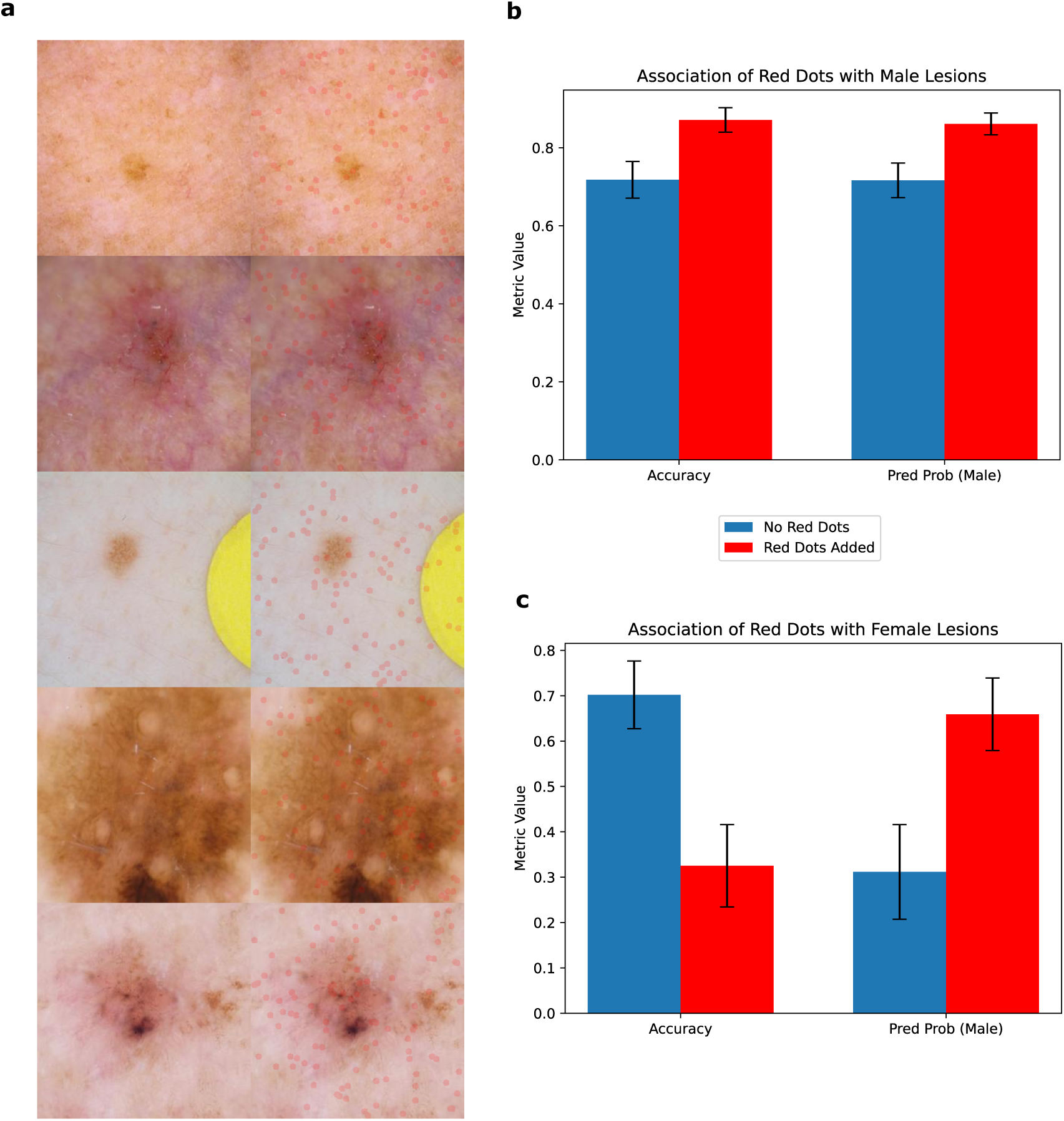
Additional experiment to verify impact of redness on sex prediction. **(a)** Sample images with redness manually added using red dots. The left image in each pair is the original real image and the right image is the manipulated one with red dots added. **(b)** Impact of adding red dots on the accuracy and predicted probability of the male lesions. When red dots are added, the accuracy and male predicted probability both increased. **(c)** Impact of adding red dots on the accuracy and predicted probability of the female lesions. When red dots were added, the accuracy decreased and the male predicted probability increased.

**Supplementary Fig. 2.**
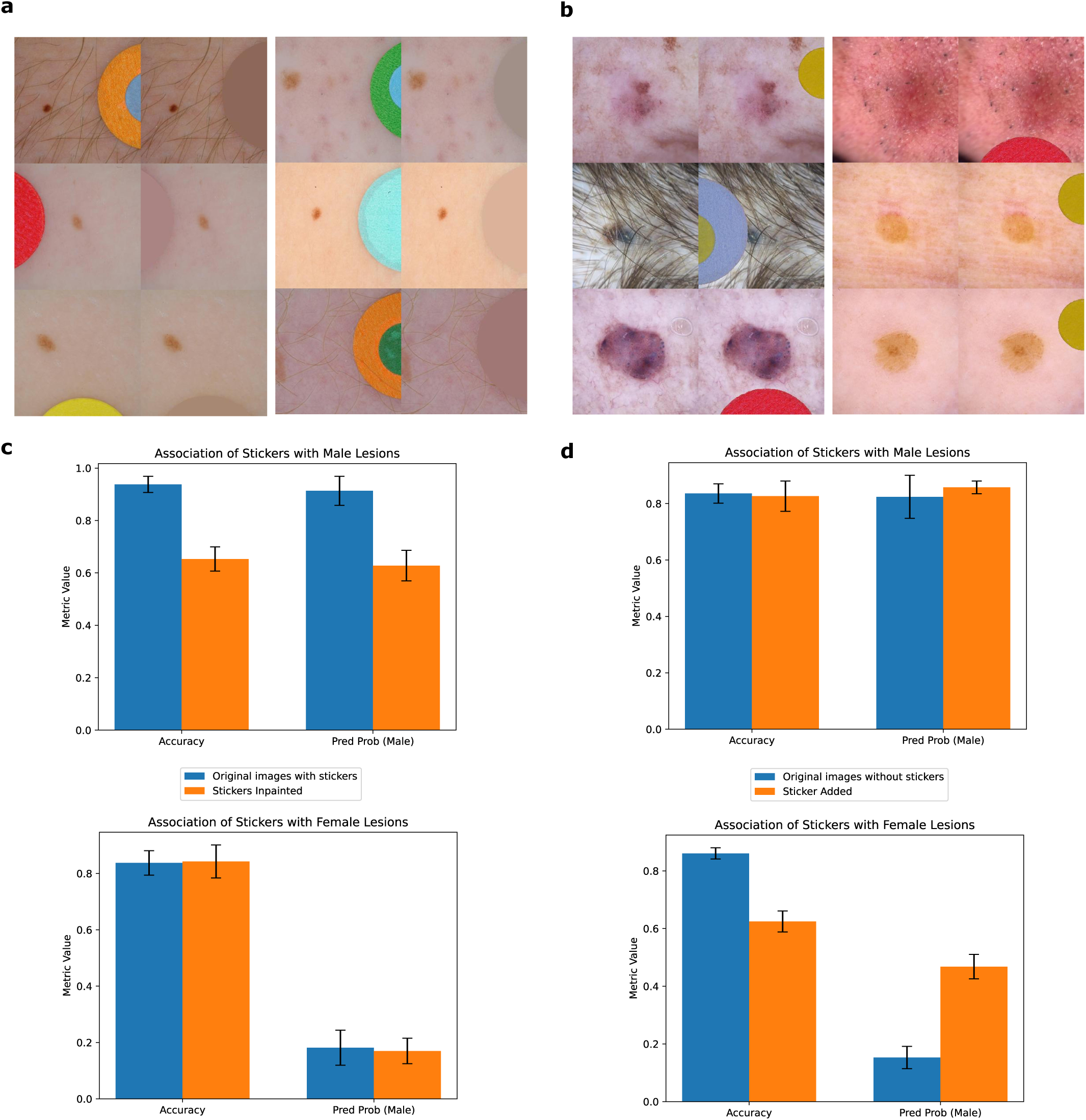
Additional experiments to verify impact of stickers on sex prediction. **(a)** Sample images with stickers manually inpainted with background color. The left image in each pair is the original real image and the right image is the manipulated one with the stickers removed using inpainting. **(b)** Sample images with stickers manually added. The left image in each pair is the original real image and the right image is the manipulated one with the stickers added. **(c)** Impact of removing the stickers by inpainting the original images (with stickers) on the accuracy and male predicted probability of the male (top) and female (bottom) lesions. When the stickers were removed, the accuracy and predicted probability of the male lesions decreased, while that of the female lesions relatively stayed the same. **(d)** Impact of adding stickers to the original images (without stickers) on the accuracy and male predicted probability of the male (top) and female (bottom) lesions. When the stickers were added, the accuracy and predicted probability of the male lesions stayed the same while for female lesions the accuracy decreased and male predicted probability increased.

**Supplementary Fig. 3.**
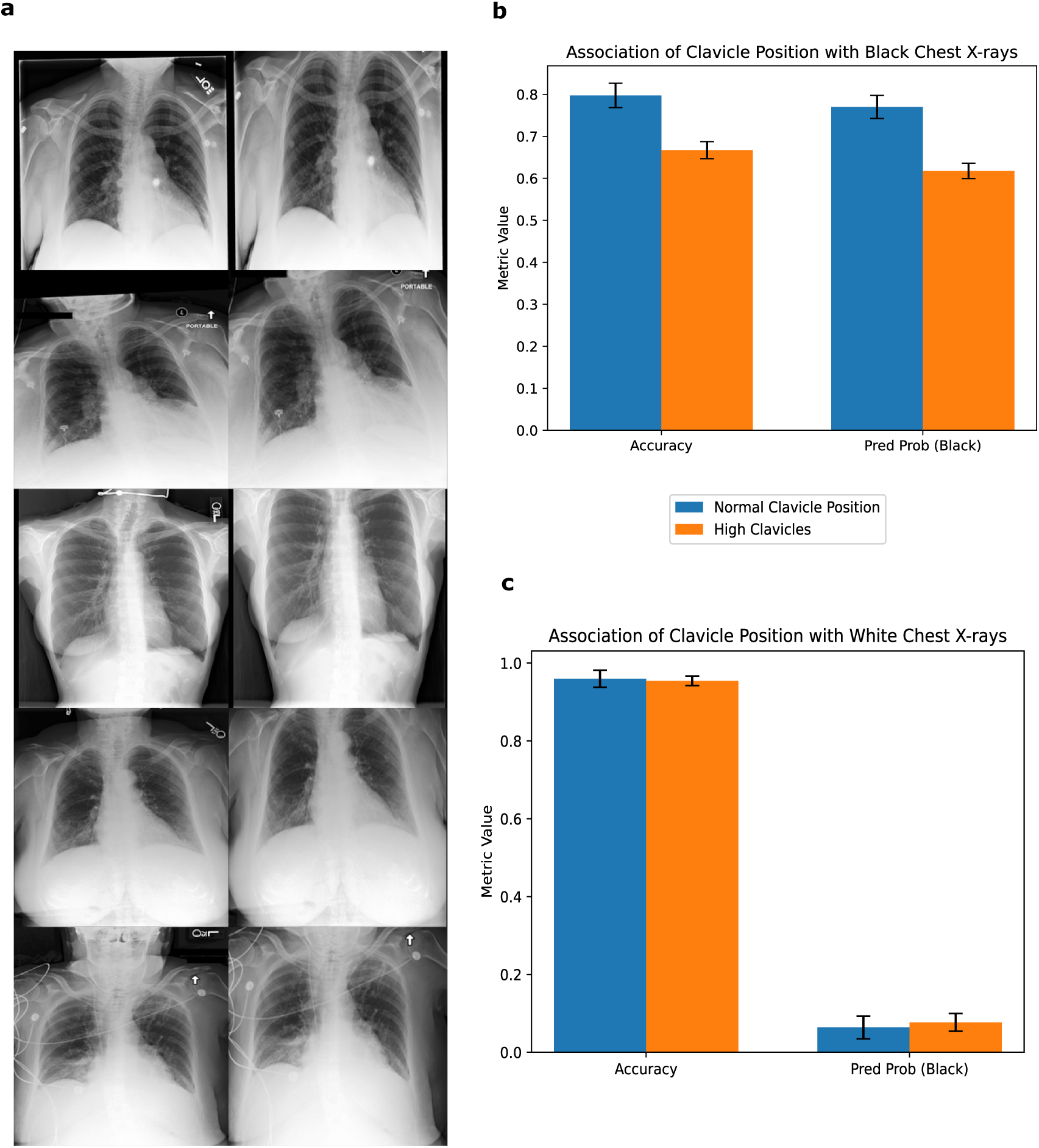
Additional experiment to verify impact of high clavicle position on race prediction. **(a)** Sample images with clavicles shifted up. The left image in each pair is the original real image and the right image is the manipulated one. **(b)** Impact of high clavicle position on the accuracy and the black predicted probability of the black chest x-rays. When the clavicles were shifted up, both the accuracy and the predicted probability decreased. **(c)** Impact of high clavicle position on the accuracy and the black predicted probability of the white chest x-rays. When the clavicles were shifted up, both the accuracy and the predicted probability stayed the same.

**Supplementary Fig. 4.**
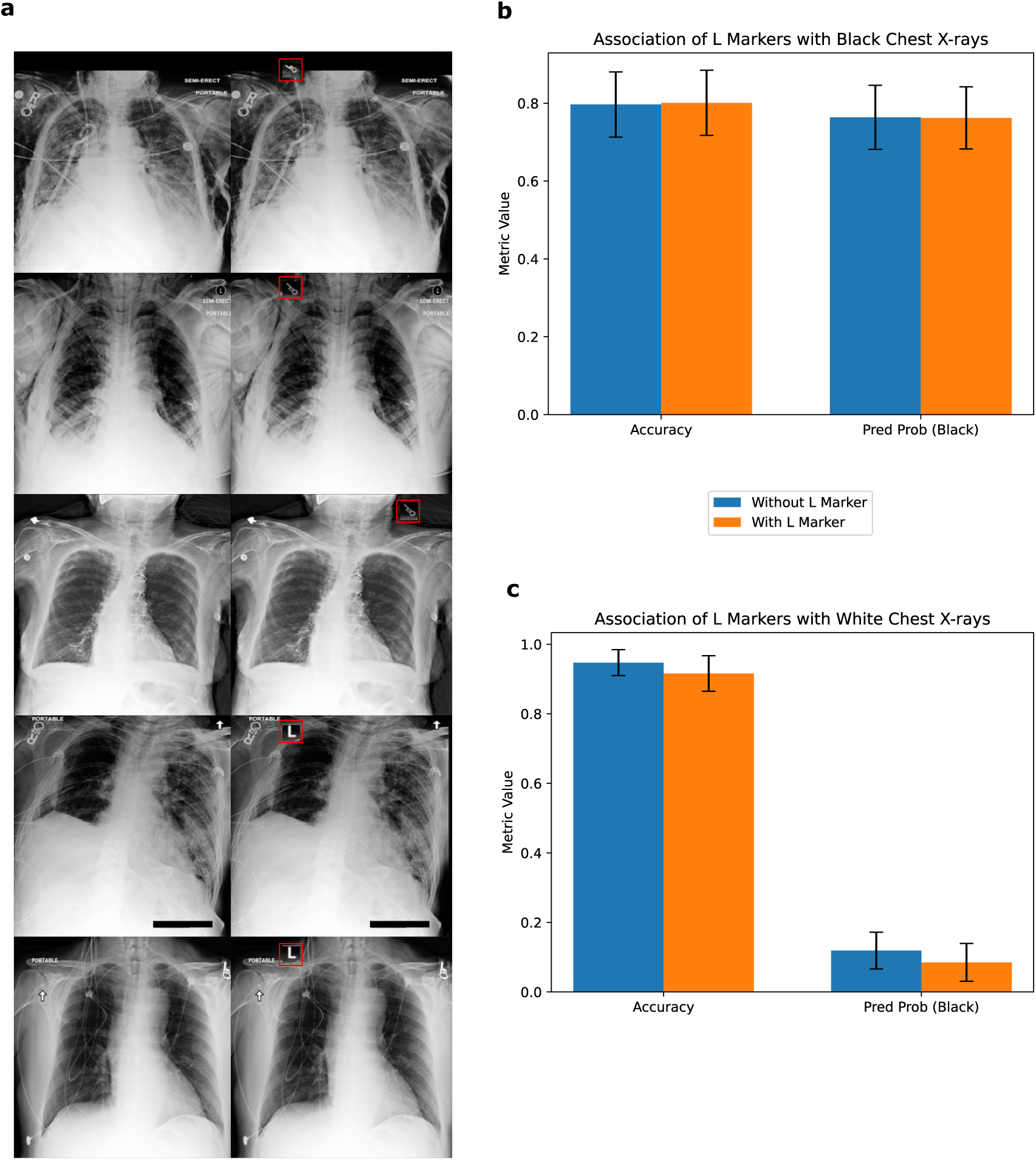
Additional experiment to verify impact of L markers on race prediction. **(a)** Sample images with L-Laterality markers added. The left image in each pair is the original real image and the right image is the manipulated one. **(b)** Impact of adding the laterality markers on the accuracy and the black predicted probability of the black chest x-rays. When the laterality markers were added, both the accuracy and the predicted probability stayed the same. **(c)** Impact of adding the laterality markers on the accuracy and the black predicted probability of the white chest x-rays. When the markers were added, both the accuracy and the predicted probability decreased slightly.

**Supplementary Fig. 5.**
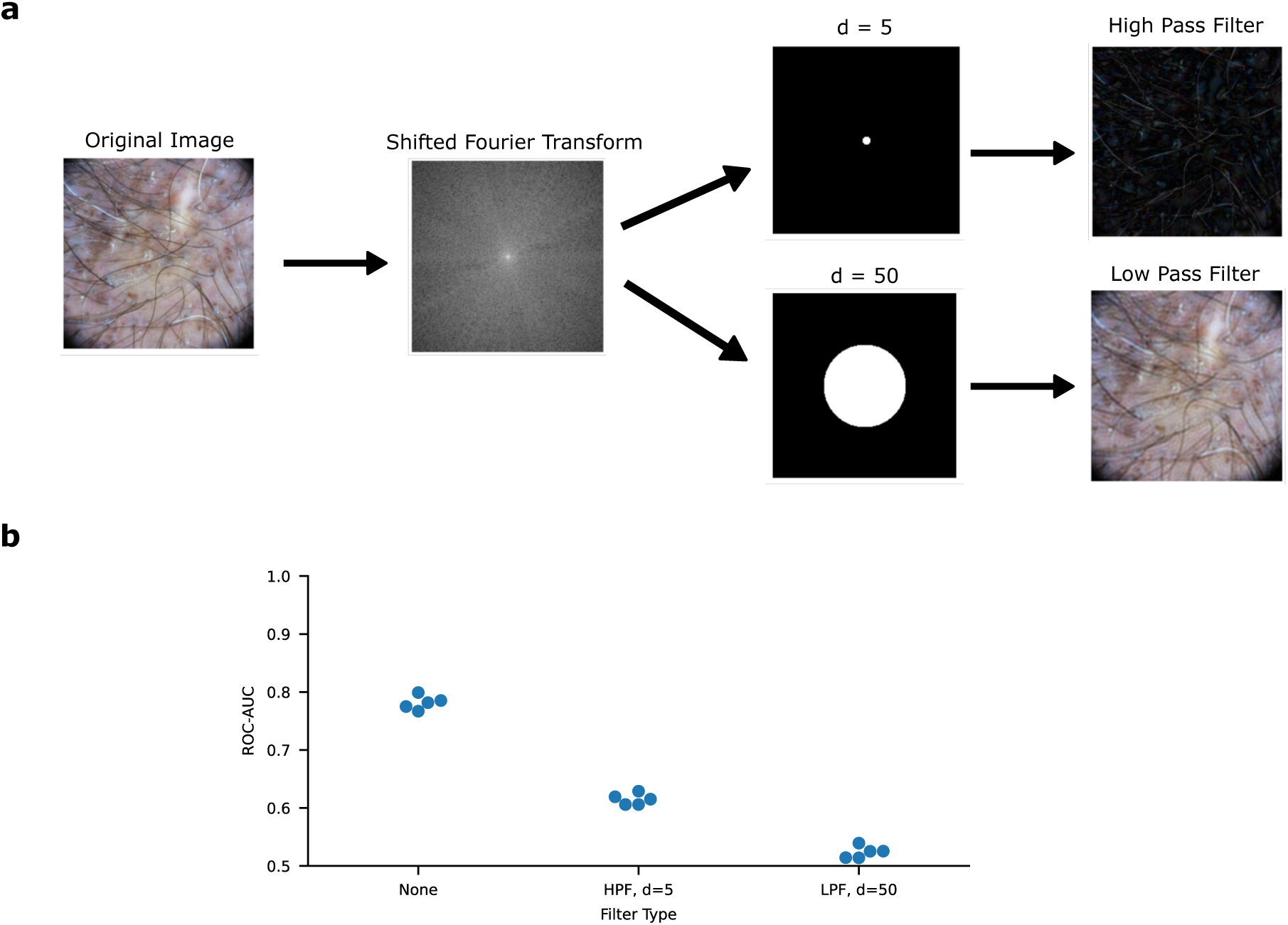
Frequency filtering experiment. **(a)** We started with a real dermoscopic image and retrieved the shifted Fourier transform to break it down into its component frequencies, with the frequencies decreasing from the center. Then, a circle mask was applied to the transform of varying pixel diameters to filter out different frequencies. For d=5, we used a high pass filter to only allow the frequencies within the circle to pass through. For d=50, we used a low pass filter to only allow for frequencies outside the circle to pass through. **(b)** Performance in terms of the ROC-AUC of using different filter types and circle masks for the sex prediction task across 5 replicates.

**Supplementary Fig. 6.**
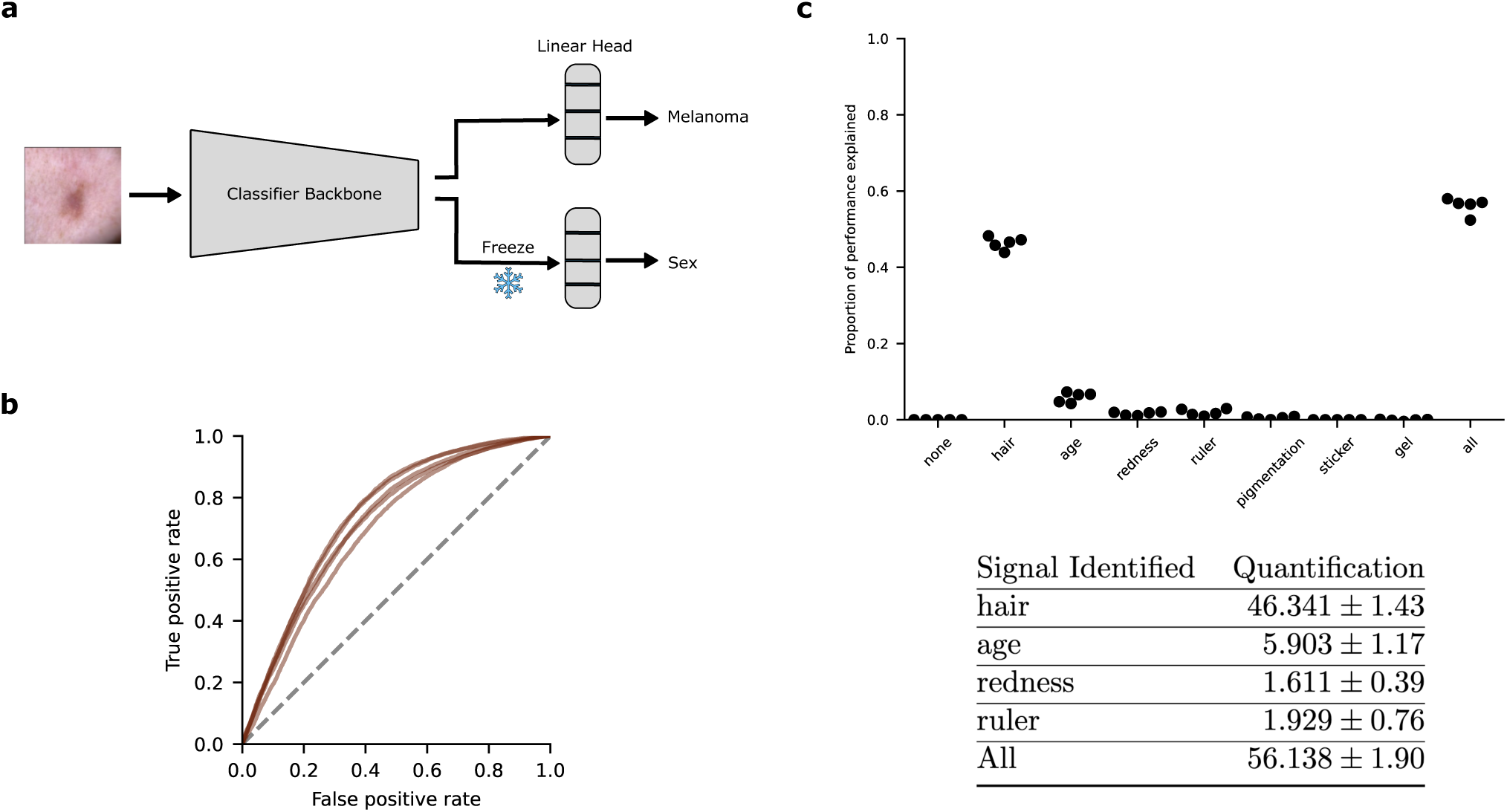
Melanoma classifier encodes demographic information. **(a)** We used transfer learning to train a sex classifier by replacing the linear head of a pre-trained melanoma classifier while keeping the backbone frozen. **(b)** Performance of training a linear classifier using a CLIP-based foundation model (MONET) embeddings for predicting sex. Each of the curves represents one of five training replicates. **(c)** Visualization of the proportion of ROC-AUC explained across five replicates along with the quantification and standard deviations.

**Supplementary Fig. 7.**
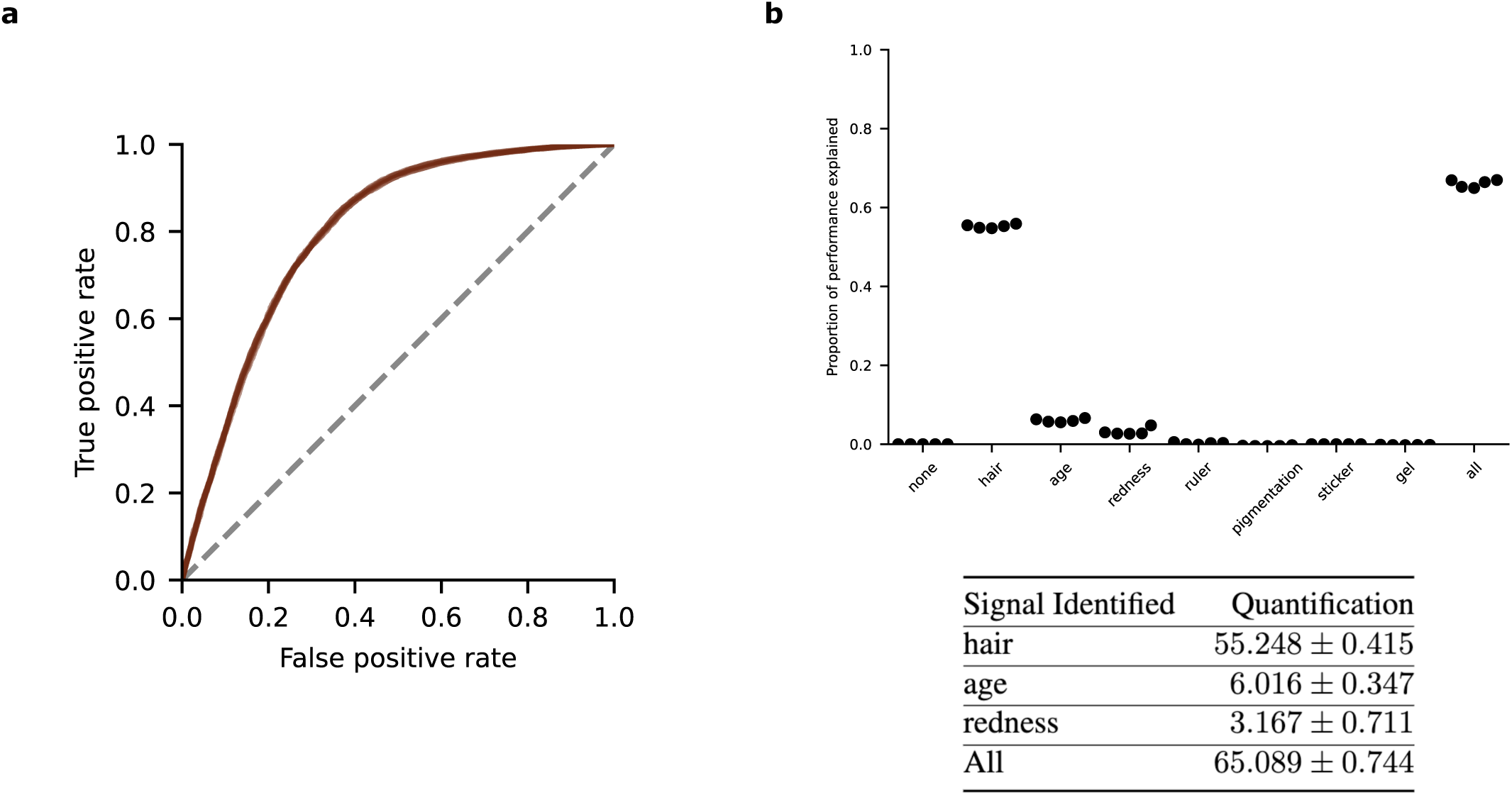
CLIP-based foundation model encodes demographic information. **(a)** Performance of training a linear classifier using a CLIP-based foundation model (MONET) embeddings for predicting sex. Each of the curves represents one of five training replicates. **(b)** Visualization of the proportion of ROC-AUC explained across five replicates along with quantification and standard deviations.

## Supplementary Tables

**Supplementary Table. 1.** Odds ratios (ORs) for predicting the female sex based on the diagnosis. Diagnoses are sorted in descending order by the absolute deviation from an OR of 1 in the training data, with those lacking at least 10 corresponding images in both the training and test data excluded. Images lacking a diagnosis were excluded from the analysis (6575 of 45924 images in the training data, and 19926 of 23461 images in the test data). N indicates the number of images.

| Lesion Diagnosis | OR, train | OR, test | N, train | N, test |
| --- | --- | --- | --- | --- |
| solar lentigo | 1.74 | 2.07 | 255 | 162 |
| lentigo simplex | 0.60 | 1.57 | 90 | 155 |
| atypical melanocytic proliferation | 1.46 | 1.07 | 13 | 86 |
| squamous cell carcinoma | 0.70 | 1.10 | 699 | 32 |
| dermatofibroma | 1.40 | 2.35 | 264 | 17 |
| basal cell carcinoma | 0.81 | 1.59 | 3278 | 21 |
| seborrheic keratosis | 0.87 | 0.55 | 1316 | 214 |
| actinic keratosis | 1.13 | 0.85 | 850 | 60 |
| nevus | 1.10 | 1.46 | 26005 | 1979 |
| melanoma | 0.99 | 0.48 | 5122 | 649 |

**Supplementary Table. 2.**
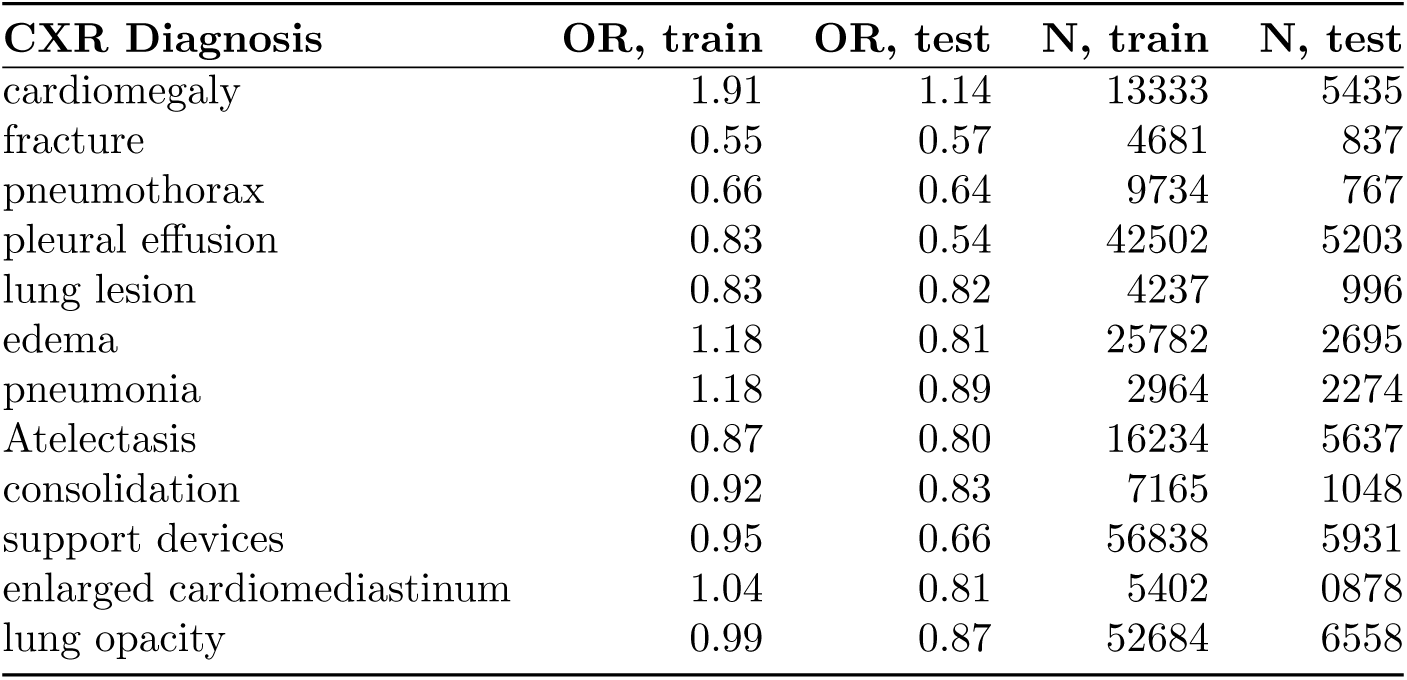
Odds ratios (ORs) for predicting the white race from chest x-rays based on the diagnosis. Diagnoses are sorted in descending order by the absolute deviation from an OR of 1 in the training data, with those lacking at least 10 corresponding images in both the training and test data excluded. N indicates the number of images.

**Supplementary Table. 3.** Odds ratios (OR) for prediction of female sex based on method of dermoscopy employed in image acquisition. Images lacking information on acquisition method are excluded (38498 of 45924 images in the training data, and 14993 of 23461 images in the test data). N indicates the number of images.

| Dermoscopy method | OR, train | OR, test | N, train | N, test |
| --- | --- | --- | --- | --- |
| contact polarized | 0.99 | 1.11 | 5277 | 459 |
| contact non-polarized | 1.03 | 0.92 | 2047 | 7963 |
| non-contact polarized | 0.77 | 0.82 | 102 | 46 |

**Supplementary Table. 4.**
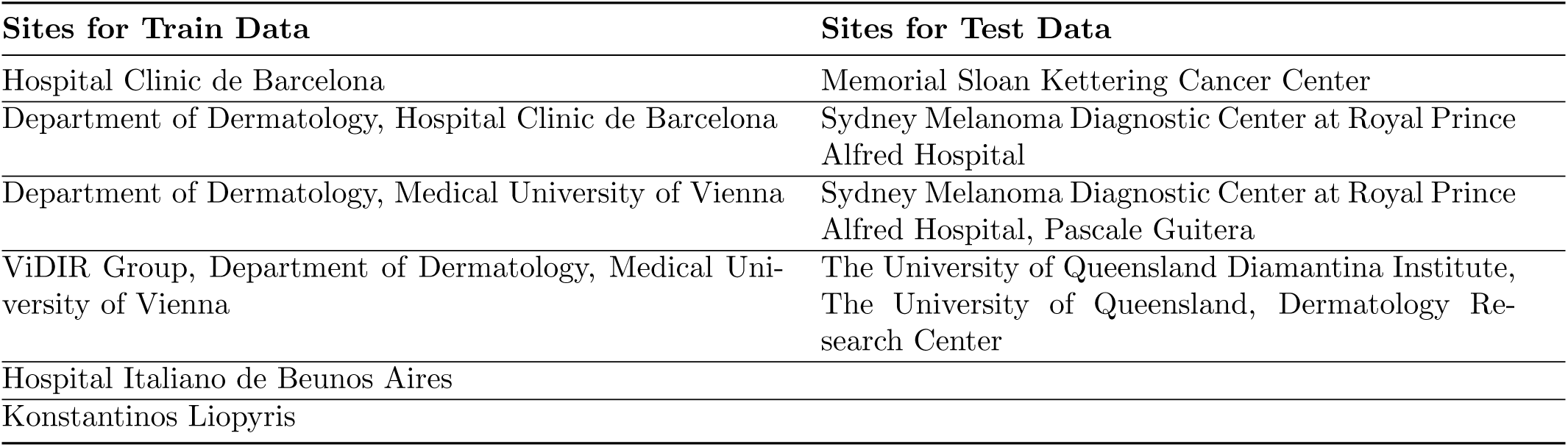
Hospital sites used in the train and test sets for the dermatology analysis.

